# Peroxisome Biogenesis Disorders in the Zellweger Spectrum: Ophthalmic Findings from a New Natural History Study Cohort and Scoping Literature Review

**DOI:** 10.1101/2022.11.06.22279732

**Authors:** Christine Yergeau, Razek Georges Coussa, Fares Antaki, Catherine Argyriou, Robert K. Koenekoop, Nancy E. Braverman

## Abstract

**Background:** Zellweger Spectrum Disorder (ZSD) is caused by bi-allelic defects in any of 13 *PEX* genes, resulting in failure to form functional peroxisomes. Individuals manifest a wide spectrum of clinical phenotypes and severity, but almost all have retinal degeneration leading to blindness. The onset, extent and progression of retinal findings has not been well-described and there are no therapies for treating vision loss. With expanding research and trials on retinal gene therapy for genetic disorders, it is now crucial to understand the natural history of vision loss in ZSD for defining reliable endpoints for upcoming interventional trials.

Here we describe ophthalmic findings in the largest number of ZSD patients to date.

**Methods:** We reviewed ophthalmology records from our retrospective longitudinal cohort of 66 patients, and cross-sectional ophthalmic findings from 79 patients reported in the literature. We divided patients by severe, intermediate or mild disease based on genotypes or their reported disease severity.

**Results:** We found that visual acuity (VA) declines slowly (+0.01 Logmar/year) with a mean of 0.93 Logmar (0 = normal vision, 1 = legal blindness) in all 53 intermediate-mild patients with available data. Longitudinal VA data revealed slow loss over time and legal blindness onset at average age 7.8 years. Fundoscopy showed retinal pigmentation, macular abnormalities, small or pale optic discs and attenuated vessels with higher prevalence in milder severity groups and did not change with age. Electroretinogram (ERG) tracings were diminished in 93% of patients, 40% of which were extinguished. ERG responses did not change over time in patients with multiple ERGs. Optical coherence tomography (OCT), reported only in milder patients, revealed cystoid macular edema or macular schisis in 16/21 (age 1.8-30 years). Serial OCTs showed evolution or stable macular edema.

**Conclusions:** Although limited by retrospective data, we highlight several useful conclusions (1) VA slowly deteriorates and is without clear association with disease severity, (2) serial ERGs are not useful for documenting vision loss progression and (3) intraretinal cysts may be common in ZSD. This study indicates that systematically reporting multiple measures will be required for accurately assessing visual function in the ZSD population, including measures of functional vision.

## Introduction

Peroxisomes are membrane-bound ubiquitous organelles required for multiple vital metabolic pathways including catabolism of very long chain fatty acids (VLCFAs) and synthesis of plasmalogens^1^. Peroxisome function and assembly depend on the coordinated action of PEX proteins, encoded by *PEX* genes. Biallelic mutations in any of 13 *PEX* genes cause Peroxisome Biogenesis Disorder (PBD) in the Zellweger Spectrum (Zellweger Spectrum Disorder, ZSD). As a result of impaired peroxisomal functions, individuals with ZSD can manifest a range of multi-systemic manifestations of varying severity, including neurological, liver, adrenal, bone involvements, hearing and vision loss^2^. Disease severity is generally associated with the level of residual PEX protein function and resulting peroxisome function^3–5^. Although the clinical presentation of ZSD is highly variable across disease severities, nearly all patients develop a progressive retinopathy leading to blindness.

Previous terminology for ZSD used the names Zellweger syndrome, neonatal adrenoleukodystrophy, infantile Refsum disease and Heimler syndrome to refer to severe, intermediate, mild and milder forms of the condition, respectively. The current nomenclature, referring to Zellweger spectrum, embraces the common etiology, the often overlapping clinical phenotypes and novel phenotypes that are being described. Patients with a severe phenotype are born with malformations that include polymicrogyria, cortical renal microcysts and chondrodysplasia punctata. Most of these individuals do not survive past the first 1-2 years of life. Patients with intermediate and milder phenotypes do not have congenital malformations, but develop progressive organ involvement due to global peroxisome dysfunction over time^6^. The majority of these patients live to adulthood and if treatment were available it could halt or ameliorate disease progression.

We are interested in potential therapies that can improve vision outcomes in intermediate and milder ZSD patients, as prevention of visual decline would have a huge impact on the life quality of these individuals and their families. However, ophthalmological findings and their rate of progression in ZSD patients have not yet been systematically characterized making it difficult to establish ideal cohorts and clear visual endpoints for clinical trials. Ophthalmic symptoms reported in severely affected patients include retinopathy, glaucoma, cataracts and corneal clouding^7^, although cataracts can also develop with age in milder patients^8,9^. Milder patients have been diagnosed with Usher Syndrome^10,11^ and usually present with retinitis pigmentosa, macular atrophy, reduced visual acuity (VA), and reduced or extinguished electroretinograms (ERGs)^12–14^. Additional descriptions include retinal arteriolar attenuation, optic nerve atrophy, nystagmus, and foveal thinning. Progressive peripheral visual field loss, nyctalopia, dyschromatopsia as well as cystoid macular edema and macular degeneration have been reported in two mild individuals^9,15^.

Our current knowledge on the occurrence and progression of ophthalmic symptoms in ZSD is based on case reports and on a few retrospective cohorts with a limited number of patients and without clear correlation between ophthalmic manifestations and overall disease severity. To address this, we present a scoping review of ophthalmic findings from 79 patients previously reported as case reports or series in the literature and from medical records of 66 patients enrolled in our retrospective natural history study on ZSD. We report both cross-sectional and longitudinal fundus descriptions, visual acuity, electrophysiological and retinal imaging findings across disease severities. Our results identify the common ophthalmic findings and their prevalence in ZSD individuals. We also describe the natural history of visual acuity and retinal structure by optical coherence tomography (OCT) exams over time. This data can be used to design a systematic prospective study on vision loss in ZSD to identify clinical endpoints and patient cohorts for future interventional trials.

## Methods

### Scoping review of literature

#### Search methods, eligibility criteria and study selection for the literature review

Studies included in this review were primary clinical research studies published in English or French that reported ophthalmic findings in patients diagnosed with ZSD. Observational studies as well as interventional studies were included for description of vision outcomes prior to the intervention(s).

We performed a comprehensive search of the medical database MEDLINE (PubMed, https://pubmed.ncbi.nlm.nih.gov/, accessed in March 2020) with each of the following terms: “Zellweger spectrum”, “Zellweger Disorder”, “Zellweger syndrome”, “Cerebrohepatorenal”, “Neonatal adrenoleukodystrophy”, “Infantile Refsum”, “Heimler”, “Peroxisome disorder” and “Peroxisome disease” in combination with each of the following terms: “Case report”, “Ocular”, “Ophthalmic”, “Vision” and “Eye”. Identified and eligible articles were collected and managed with the EndNote and Excel softwares. Search results were screened (CA, CY, NEB) by title and abstract and selected full-text articles were reviewed for eligibility. Inclusion criteria for this review were studies that reported any ophthalmic or vision findings in patients diagnosed with ZSD. Studies that did not report ophthalmic or vision findings in patients with ZSD or that reported ophthalmic or vision findings in other diseases were excluded. Due to our career focus and expertise in ZSD, we collect articles related to peroxisome disease and disorders from the existing literature. We also searched this collection and identified additional articles in which ophthalmic findings in ZSD patients were described in the text but did not come out in the PubMed search.

#### Data collection and synthesis for the literature review

The following data were collected by (FA) and reviewed (CY) from the selected articles, if available: the patient’s age at the initial retinal exam (presumed to be the age of retinopathy diagnosis unless age at diagnosis was explicitly and separately specified in the article), sex, genotype, VA of both eyes if available, refractive error of both eyes, presence of nystagmus, strabismus, anterior segment exam findings of both eyes, fundus findings of both eyes, description of visual field, relevant ophthalmic imaging including optical coherence tomography, electroretinographic studies, as well as any other reported eye or vision-related findings. Reporting bias was assessed to account for differences between reported and unreported findings. To minimize the reporting bias associated with the differences between reported and unreported findings across studies, we interpreted the unreported findings as unavailable instead of as negative findings. Data was collected using Excel software. The data was organized in a data synthesis matrix to include all collected data for each article (Supplementary Table 5). Then, the collected data was organized in a second data synthesis matrix to separate the findings per individual patient to allow for quantification of patients with specific findings (Supplementary Table 4).

#### Natural History Study - Patient Recruitment & Medical Chart Data Collection

We collected medical records from 66 individuals with ZSD from the United States, Canada, Europe and Australia enrolled in our Longitudinal Natural History Study on peroxisomal disorders at the McGill University Health Center (MUHC) from January 2012 to January 2019. This retrospective study adheres to the tenets of the Declaration of Helsinki, was approved by the MUHC review ethics board (Study #11-090-PED) and is registered and publicly available on clinicaltrials.gov (Identifier: NCT01668186). Individuals enrolled in this study already received a diagnosis of ZSD based on molecular and biochemical testing at their local health care centers and were included in this study after obtaining informed consent. We obtained medical records from each participant’s local health care institution after we received the authorization from the patient or the parent/legal representative. We reviewed the medical records and input relevant data on ophthalmic findings into our custom-made Microsoft Access database. Clinical data extracted included genetic testing results, visual acuity, refraction, nystagmus assessment, slit-lamp and fundoscopy examination findings, OCT findings and ERG results. Clinical information was obtained at baseline as well as during follow-ups visits for each patient. It is important to note that some clinical data was not available for each patient at every follow-up visit.

#### Disease Spectrum Staging

To standardize the genotypic and phenotypic subgroups among the literature patients and our natural history study cohort, we grouped the patients into four different severity categories based on specific genotypes or clinical phenotypes. Genotypes were available in all 66 patients in our natural history study cohort and in 36 out of 79 patients from the literature. First, if patients were known homozygotes for the common hypomorphic allele *PEX1* c.2528G>A (p.Gly843Asp), which is associated with a mild ZSD phenotype^3,5^, we grouped them into the “PEX1 p.G843D/G843D - Mild ZSD” group. Then, if patients were known compound heterozygotes for PEX1 p.G843D and a non-functional allele, we grouped them into the “PEX1 p.G843D/Null - Intermediate ZSD “ group. The most common non-functional allele for patients in this group is the common *PEX1* p.Ile700Tyrfs42X null variant (11/20 patients in this severity group)^16,17^. This genotype-phenotype correlation for the common PEX1-p.G843D allele in homozygosity as a mild phenotype and “hemizygosity” as an intermediate phenotype has been a consistent reported observation^3,5,6,13,18,19^. The majority (70%) of all ZSD patients have mutations in *PEX1*, and p.G843D represents 30% of all *PEX1* alleles.

Remaining ZSD patients whose genotypes were different than those of the two first groups or unavailable were classified in two other categories based solely on their overall clinical presentation^20^. The “Severe ZSD” category includes patients who presented with the classic Zellweger Syndrome clinical phenotype with the following symptoms: seizures at birth, brain polymicrogyria on magnetic resonance imaging, renal cortical cysts on ultrasound, chondrodysplasia punctata in hips and/or knees on X-rays, as well as failure to thrive and hypotonia^21,22^. If the clinical phenotype was not well described, those who were diagnosed with “Cerebrohepatorenal syndrome” or “Zellweger syndrome” were included in the “Severe ZSD” category^6,22^. Finally, the “Mild-intermediate ZSD” category included all other patients who do not belong to the first three groups. Patients from the literature whose clinical phenotypes were not well described and that were given the diagnosis of “Heimler syndrome”^14,23^, “Infantile Refsum’s disease” or “Neonatal adrenoleukodystrophy”^6,23^ were included in the “Mild-intermediate ZSD” category.

#### Data analyses

The Snellen VA for each eye from each patient (both literature and our cohort) was converted to logMAR equivalent. As previously described by Roberts *et al*.^24^, the following logMAR values: 2.6, 2.7, 2.8 and 2.9, were used for VA recorded as counting fingers, hand movement, light perception, and no light perception, respectively. Patients without VA in both eyes were excluded from the analysis. Similarly, patients with visual assessment of “fix and follow” as well as “central/steady/maintained (CSM)” were also excluded due to the absence of a standardized Snellen equivalent. Interocular VA differences between the right and left eyes were compared using paired t-tests. For the purpose of the analysis, the VA of the right and left eye for each patient was averaged in order to account for disease asymmetry and inter-individual pathologic differences. A cross-sectional analysis in which the mean logMAR VA values were plotted against their respective age was undertaken. The significance of the correlation was tested using a linear regression model. Similarly, for our patients with follow-up data for at least 6 months apart, we compared their initial and final logMAR VA using t-tests. We also plotted the difference between their final and initial logMAR VA against their respective initial age, and we tested the latter correlation using a linear regression model. For each of our patients with follow-up data for at least 6 months apart, we have also computed linear regression analysis of the mean logMAR VA values and respective age. We have performed VA-disease severity correlations on the literature patients and our cohort by one-way ANOVA. Finally, prevalence of ophthalmic symptoms in both patients from our natural history study cohort and the literature were compared across the severity groups and tested for significant difference with the Fisher’s two-sided exact test.

#### Statistical analyses

Microsoft Excel (Microsoft Corporation, Redmond, Washington) and IBM SPSS Statistics (version 26, IBM Corporation, Armonk, New York) were used for the statistical analysis. Statistical significance was set at p<0.05 for all comparisons.

## Results

### Literature Search Results

After removing studies that did not meet the inclusion criteria or were duplicates, our PubMed search yielded 31 full-text articles that were included in this review.

### Distribution of genotypes and disease severity

Genotypes, sex and ages at ophthalmic findings for each patient in our retrospective cohort are shown in Supplementary Table 3. Most patients had mutations in the *PEX1* gene (n = 47, 70%), with most patients within this group having at least one PEX1-p.G843D allele (n = 35, 74%) which is found in homozygosity in 20 patients (57%) and in the compound heterozygous state in 20 individuals, most often paired with the null PEX1-p.I700Yfs42X allele (n = 12, 60%). Patients in the Mild-Intermediate ZSD group (n = 27) had mutations in *PEX1, PEX5, PEX6, PEX12, PEX16, PEX19* and *PEX26* genes, with the majority of patients having mutations in *PEX1* (n = 11, 41%) and in *PEX6* genes (n = 9, 33%). Amongst patients with *PEX1* and *PEX6* defects, 9 had two missense mutations in one *PEX* gene (33%) and 8 patients had one missense mutation paired with a non-functional allele in one *PEX* gene (30%).

From the literature, genotypes were available in 36 out of the 79 patients. 14/36 (39%) patients with available genotypes were PEX1-p.G843D homozygotes, 20/36 (56%) patients had variants resulting in missense amino acid substitutions (73%), premature stop codons (8%), splice site variants (5%) and deletions and duplications (14%) in *PEX1, PEX11*β, *PEX6* and *PEX26*, and 2/36 patients were *PEX1* compound heterozygotes for p.G843D and p.I700Yfs42X. From the literature, males and females were equally represented (49% and 51%, respectively), while our retrospective cohort was composed of 61% (n = 40) of males and 39% (n = 26) of females (Supplementary Table 4).

### Ophthalmic Clinical Findings

Ophthalmic findings for patients from our retrospective cohort and from the literature are summarized in Figure 1. 50% of patients in our cohort and 35% of literature patients are legally blind (visual acuity ≥ 1.00 LogMAR) without clear association between prevalence of poor visual acuity and disease severity. Most patients had nystagmus (76% in our cohort and 57% in the literature), with higher prevalence in milder severity groups only for our cohort (25% of patients in the Severe ZSD group compared to 88%, 73% and 80% of patients in the PEX1 p.G843D/Null -Intermediate ZSD, Mild-Intermediate ZSD and PEX1 p.G843D/G843D – Mild ZSD groups, respectively). Cataracts were reported in 57% of patients in our cohort and in 16% of the literature patients, without significant association with disease severity. Fundus examination results revealed that abnormal pigment spots in peripheral retina is the most common findings reported and is found in all patients in the PEX1 p.G843D/G843D - Mild group from our cohort and the literature, while it is ranging from 74% to 64% of patients from Severe to Mild-Intermediate groups. Retinal degeneration is reported in 28% of all patients combined without significant association with disease severity. Additional ophthalmic findings and age ranges at examination are provided in Supplementary Tables 1 and 2. Other fundus findings reported included macular abnormalities (34% of all patients in our cohort with higher prevalence in the PEX1 p.G843D/G843D - Mild ZSD group (73%), and 25% of literature patients without significant association with disease severity), small and/or pale optic discs (46% of all patients in our cohort with higher prevalence in the PEX1 p.G843D/G843D - Mild ZSD group (67%), and 13% of literature patients without significant association with disease severity) and attenuated retinal vessels (32% of all patients in our cohort and 21% of literature patients, without significant association with disease severity) (Supplementary Tables 1 and 2). Refraction data was available for 48 patients in our retrospective cohort, 38 of which (80%) were hyperopic (mean refractive error = 4.10) and none were myopic. All patients with at least one PEX1 p.G843D allele were hyperopic (n = 30) (data not shown).

**Figure 1.**
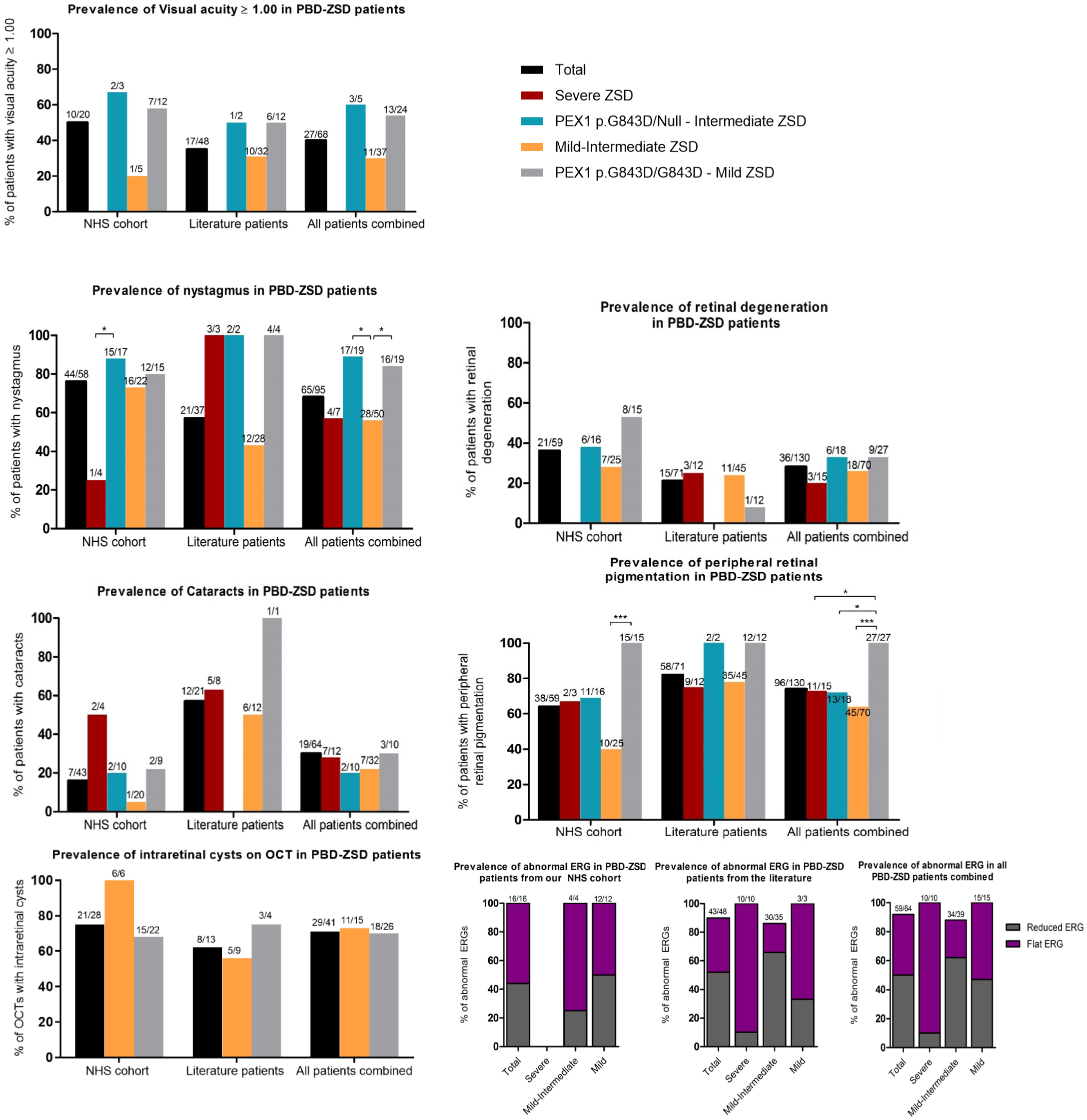
Prevalence of ophthalmic findings in ZSD patients from our retrospective natural history study (NHS) cohort and from the literature. Bars show percentage of patients with a specific ophthalmic finding. Top of bar: number of patients with findings out of total patients with assessment; colored bars represent disease severity subgroups; black bars show prevalence for patients of all severities combined. Visual acuity results were converted to Logarithm of the Minimum Angle of Resolution (logMAR). Fisher’s two-sided exact test was performed for all data to test whether the likelihood of having a specific symptom is significantly different between severity groups. Comparisons were done between Severe and Intermediate groups, Intermediate and Mild-Intermediate groups, Mild-Intermediate and Mild groups, and between Severe and Mild groups. Statistically significant differences are shown with brackets (p < 0.001 (***) and p < 0.05 (*)). ERG: Electroretinogram, OCT: Optical Coherence Tomography.

In our retrospective cohort, we estimated the age at onset of vision loss by reporting the earliest ages at which ophthalmological exam results were available. There are patients with first exams as early as one month of age across all severity groups. The mean age at first exam decreases with disease severity (17.1 years in the PEX1 p.G843D/Null - Intermediate ZSD group, 8 years in the Mild-Intermediate ZSD group (note that there are six patients with normal vision in this group) and 18.9 years in the PEX1 p.G843D/G843D - Mild ZSD group). We found that the first age at which the fundus abnormalities mentioned above are reported is not associated with overall disease severity in all three Intermediate to Mild groups (the age ranges are narrower in the severe groups due to short lifespan). We found that once a fundus abnormality is reported, it does not change with age for individual patients (Supplementary Table 3).

### Visual Acuity

A total of 20 patients from our cohort and 33 patients from the literature review were included (Table 1). For patients in our cohort, the mean age was 5.8 years old (median:4.3) and the mean logMAR VA was 0.79 years old (median: 0.79, range: 0.30 to 1.60). For literature review patients, the mean age was 18.9 years old (median: 18.5, range: 4.0 to 35.0) and the mean logMAR VA was 0.82 (median: 0.80, range: 0.00 to 1.55). The mean age was 14.0 yo (median: 12.0, range: 1 to 35.0) and the mean logMAR VA was 0.81 years old (median: 0.80, range: 0.00 to 1.60) for both groups combined.

**Table 1.** Mean logMAR visual acuity (VA) with age ranges for previously reported patients as well as patients in our retrospective natural history study cohort with Zellweger Spectrum Disorder per disease severity subgroup. VA from our retrospective cohort patients are shown in white and VA from literature patients are shown in light blue. Abbreviations: VA = Visual acuity.

| **Disease severity subgroup** | **PEX1 p.G843D/Null – Intermediate ZSD** |  | **Mild – Intermediate ZSD** |  | **PEX1 p.G843D/G843D – Mild ZSD** |  |
| --- | --- | --- | --- | --- | --- | --- |
|  | VA (logMAR) | Age range (y) | VA (logMAR) | Age range (y) | VA (logMAR) | Age range (y) |
|  | 0.70 | 0 - 4 | 0.85 | 0 - 4 | 0.95 | 0 - 4 |
|  | 0.90 | 0 - 4 | 0.65 | 0 - 4 | 0.60 | 0 - 4 |
|  | 1.60 | 0 - 4 | 0.88 | 5 - 9 | 0.55 | 0 - 4 |
|  | 1.3 | 15 - 19 | 1.05 | 10 - 14 | 0.85 | 0 - 4 |
|  |  |  | 0.73 | 10 - 14 | 0.40 | 0 - 4 |
|  |  |  | 0.5 | 0 - 4 | 0.45 | 0 - 4 |
|  |  |  | 0.2 | 0 - 4 | 0.30 | 5 - 9 |
|  |  |  | 0.5 | 5 - 9 | 0.78 | 5 - 9 |
|  |  |  | 1 | 5 - 9 | 0.80 | 5 - 9 |
|  |  |  | 0.25 | 5 - 9 | 0.75 | 5 - 9 |
|  |  |  | 0 | 5 - 9 | 1.00 | 10 - 14 |
|  |  |  | 1.55 | 10 - 14 | 1.00 | 15 - 19 |
|  |  |  | 1 | 10 - 14 | 0.69 | 5 - 9 |
|  |  |  | 1 | 15 - 19 | 0.7 | 15 - 19 |
|  |  |  | 0.6 | 15 - 19 | 0.6 | 15 - 19 |
|  |  |  | 0.8 | 15 - 19 | 0.6 | 15 - 19 |
|  |  |  | 1 | 15 - 19 | 1 | 20 - 24 |
|  |  |  | 1 | 15 - 19 | 1.4 | 20 - 24 |
|  |  |  | 0.45 | 15 - 19 | 1.3 | 25 - 29 |
|  |  |  | 1 | 20 - 24 | 1.3 | 25 - 29 |
|  |  |  | 1 | 20 - 24 | 0.65 | 25 - 29 |
|  |  |  | 1 | 25 - 29 | 1.3 | 30 - 34 |
|  |  |  | 0.65 | 25 - 29 | 1.1 | 30 - 34 |
|  |  |  | 0.7 | 25 - 29 | 0.5 | 35 - 39 |
|  |  |  | 0.45 | 30 - 34 |  |  |
| **Mean** | 1.13 | 6.0 | 0.77 | 14.6 | 0.82 | 15.1 |
| **Median** | 1.10 | 2.7 | 0.83 | 13.5 | 0.77 | 15.2 |
| **Count** |  | 4 |  | 25 |  | 24 |
| **% of cohort** |  | 7.55 |  | 47.17 |  | 45.28 |

### Cross-sectional Analysis

The linear regression for the logMAR VA vs. age for our patients was not statistically significant (Figure 2, *p=*0.3), while that for the literature patients showed a statistically significant slow but progressive decline with increasing age (Figure 2, *p=*0.03). Similarly, the linear regression for the logMAR VA vs. age for all patients was statistically significant (Figure 2, *p=*0.03). Specifically, over a 10 years period, the VA drop was estimated at 0.1 logMAR, which is equivalent to ∼ one line on the Snellen chart.

**Figure 2.**
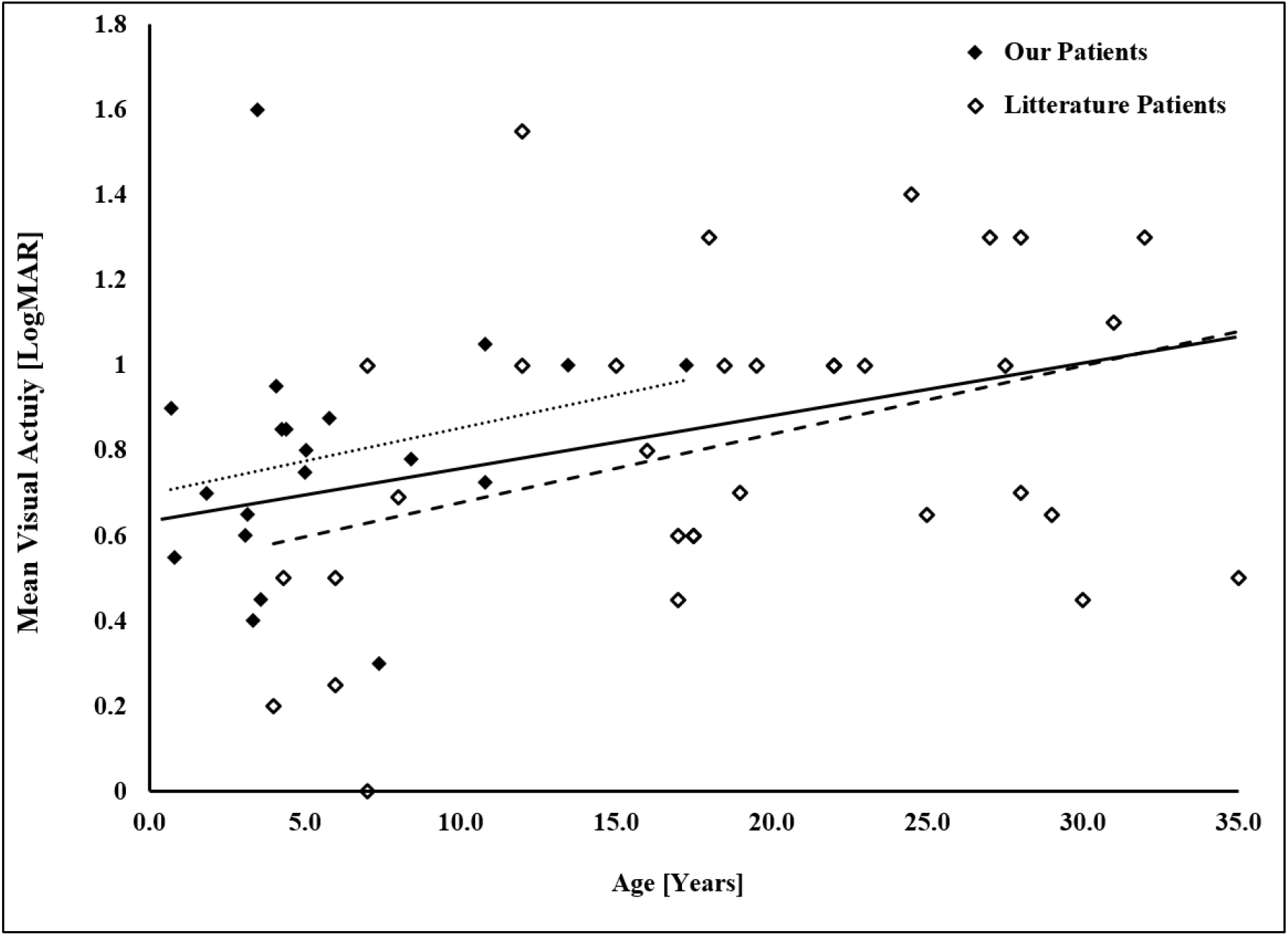
Mean logMAR visual acuity (VA) as a function of age for previously reported patients as well as patients in our retrospective cohort with Zellweger Spectrum Disorder. The linear regression for the logMAR VA vs. age for our patients (N= 20) was not statistically significant (pointed line: y = 0.015x + 0.70, *p=*0.3). The linear regression for the logMAR VA vs. age for the literature patients (N= 33) was statistically significant (dashed line: y = 0.016x + 0.52, *p=*0.03). The linear regression for the logMAR VA vs. age for all patients (N= 53) was statistically significant (solid line: y = 0.010x + 0.67, *p=*0.03).

### Genotype-phenotype correlations

Grouping the patients from the retrospective cohort and from the literature into our disease severity subgroups yielded 24 patients (45.3%) in the PEX1 p.G843D/G843D - Mild ZSD group, 25 patients (47.2%) in the Mild-Intermediate ZSD group and 4 patients (7.5%) in the PEX1 p.G843D/Null - Intermediate ZSD group for the visual acuity analysis (Table 1). In the PEX1 p.G843D/G843D - Mild ZSD group, the mean age was 15.1 years (median:15.2) and the mean logMAR VA was 0.82 years (median: 0.77, range: 0.30 to 1.40). In the Mild-Intermediate ZSD group, the mean age was 14.6 years (median:13.5) and the mean logMAR VA was 0.77 years (median: 0.83, range: 0.00 to 1.55). In the PEX1 p.G843D/Null - Intermediate ZSD group, the mean age 6.0 years (median: 2.7) and the mean logMAR VA was 1.13 years (median: 1.10, range: 0.70 to 1.60). There were no statistically significant differences in one-way ANOVA comparing the ages among the 3 severity groups (*p=*0.23) as well as the logMAR VA (*p=*0.12). However, patients in the PEX1 p.G843D/Null - Intermediate ZSD group did present earlier and had a worse logMAR compared to both those in the PEX1 p.G843D/G843D - Mild ZSD and Mild-Intermediate ZSD groups (Table 1).

### Longitudinal analysis

Fourteen patients in our cohort had longitudinal data with a mean follow-up period of 8.4 years (median: 6.4, range: 2.3 to 27.0, Supplementary Table 3). In the PEX1 p.G843D/G843D - Mild ZSD group, the mean age was 11.2 years (median: 9.3) and the mean logMAR VA was 0.90 (median: 0.90, range: 0.3 to 2.0). In the Mild-Intermediate ZSD group, the mean age was 7.1 years (median:7.5) and the mean logMAR VA was 0.80 (median: 0.9, range: 0.70 to 0.9). In the Intermediate ZSD group, the mean age was 5.5 years (median: 4.6) and the mean logMAR VA was 1.30 (median: 1.30, range: 0.7 to 1.60). Figure 3 shows the spread of VA progression over time for each patient and per severity categories. Despite the scattering of data points, especially in the PEX1 p.G843D/G843D - Mild ZSD group, there seems to be a dichotomous distinction in which patients in the Intermediate ZSD group present with poor VA at an earlier age compared to those in the other two groups. Despite their statistical non-significance, the linear regressions of the logMAR VA vs. age for each group showed a very slow decline over time (Figure 3).

**Figure 3.**
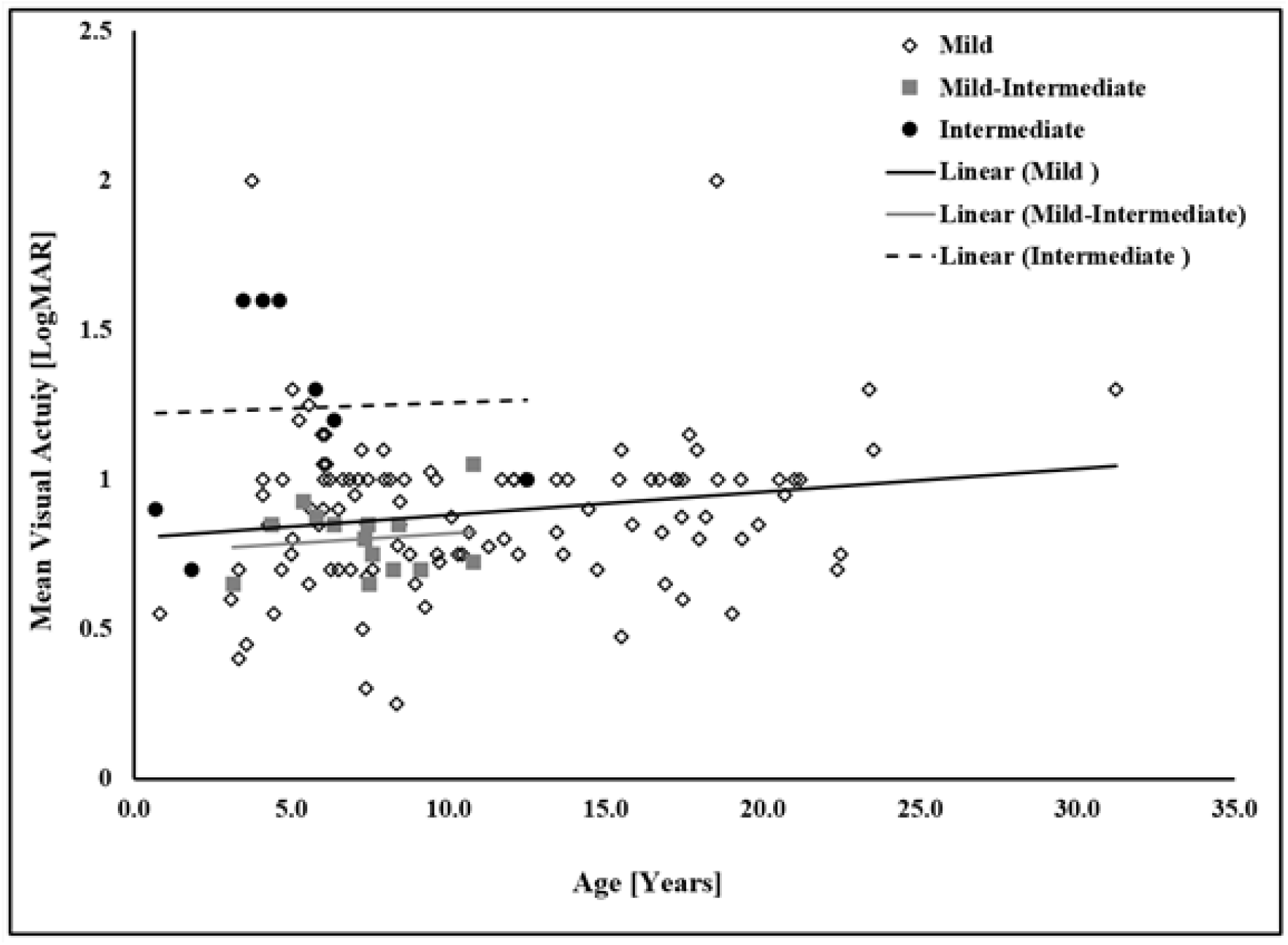
Longitudinal logMAR visual acuity as a function of age for patients in our retrospective cohort with Zellweger Spectrum Disorder grouped by disease severity. The linear regression of the logMAR VA vs. age in the PEX1 p.G843D/G843D – Mild ZSD (“Mild”) group (n= 106) was not statistically significant (solid black line: y = 0.008x + 0.81, *p=*0.06). The linear regression of the logMAR VA vs. age in the Mild-intermediate ZSD (“Mild-Intermediate”) group (n= 11) was not statistically significant (grey solid line: y = 0.007x + 0.76, *p=*0.64). The linear regression of the logMAR VA vs. age in the PEX1 p.G843D/Null – Intermediate ZSD (“Intermediate”) group (n= 9) was also not statistically significant (dashed black line: y = 0.004x + 1.22, *p=*0.92).

### Electrophysiological Studies

ERG findings for patients in our cohort and from the literature are summarized in Figure 1. All ERGs done in patients from our cohort ranging from infancy to their third decade were abnormal (n = 16), with 56% showing no detectable photopic and scotopic responses and 44% showing diminished responses. 43/48 ERGs reported in the literature were abnormal, 42% and 58% of which showed no detectable responses and diminished responses, respectively. We did not find that the extent of ERG abnormalities was associated with disease severity in our cohort, however extinguished ERGs are more common in severe patients (90%) compared to Mild-Intermediate ZSD (23%) to PEX1 p.G843D/G843D - Mild ZSD patients (66%) in the literature.

Serial ERG studies in mild patients of our cohort revealed that the a- and b-wave amplitudes do not vary with age in these individual patients (data not shown). Visual evoked potential (VEP) studies were available in 4 patients in our cohort ranging from infancy to their third decade, one of which was normal, and the three others had decreased amplitude and/or delayed latency of the P100 waveform (Supplementary Table 1). 16/28 VEP studies from the literature show abnormal tracings with all 7 Severe ZSD patients and 9/21 Mild-Intermediate ZSD patients showing decreased amplitude and/or delayed latency (there were no VEPs reported in the two other severity subgroups) (Supplementary Table 2).

### Retinal Imaging

We reviewed findings from 28 OCT studies from 8 patients in our cohort and from 13 OCT studies previously reported. 7/8 patients in our cohort with OCT data available had schitic changes (in 21 out of the 28 OCT data/images collected), while schitic changes were reported in 8/13 patients from the literature. Presence of intraretinal cysts was not associated with disease severity in either or both groups combined (Figure 1). Prevalence of specific features from OCTs of patients from the literature and our cohort are summarized in Figure 1 and in Supplementary Tables 1 and 2. Written descriptions of individual OCT findings from the literature and from our cohort are found in Supplementary Tables 3 and 4, respectively.

We found in OCTs from patients in our cohort that most patients showed outer nuclear layer (ONL) schitic changes ranging from mild (Figure 4 B-C), to moderate (Figure 5 E-J) to severe (Figures 6, 7 C-D, 7 G-H, and 8 E-J). Most patients with documented OCT imaging showed ONL schitic changes in their first and second decades of life (Figures 4, 6, 7, 8 and 9). These schitic changes were seen as early as between 0 and 4 years old (Figure 6A). Additionally, the schitic changes evolve over time and can either regress (Figure 8) or increase in size (Figures 6 and 7). One patient showed absence of schisis, which was replaced by marked macular atrophy with overlying pigmentary changes in her third decade (Figure 9). Most patients showed nummular hypopigmented spots throughout their fundus (Figures 4 D-E, 5 A-D, 8 C-D and 9 A-B). There were round deep hyper-pigmented spots seen in the periphery (Figure 6A-B), mid-periphery (Figures 7 A-B, and E-F) but also extending posteriorly to the vascular arcades of the posterior pole (Figure 8 C-D). Patient 5 and patient 60 in our cohort received carbonic anhydrase inhibitor (CAI) eye drop treatments for five months and six months respectively. OCT images for patient 5 shows larger swelling of the fovea at 8 months after treatment and onwards (Figure 6). OCT reports for patient 60 describe stable macular edema for his first four OCT exams in the span of 10 months and less macular edema 10 months after the treatment stopped (Supplementary Table 3). Both patients stopped the treatment due to no improvement noted by their ophthalmologists.

**Figure 4.**
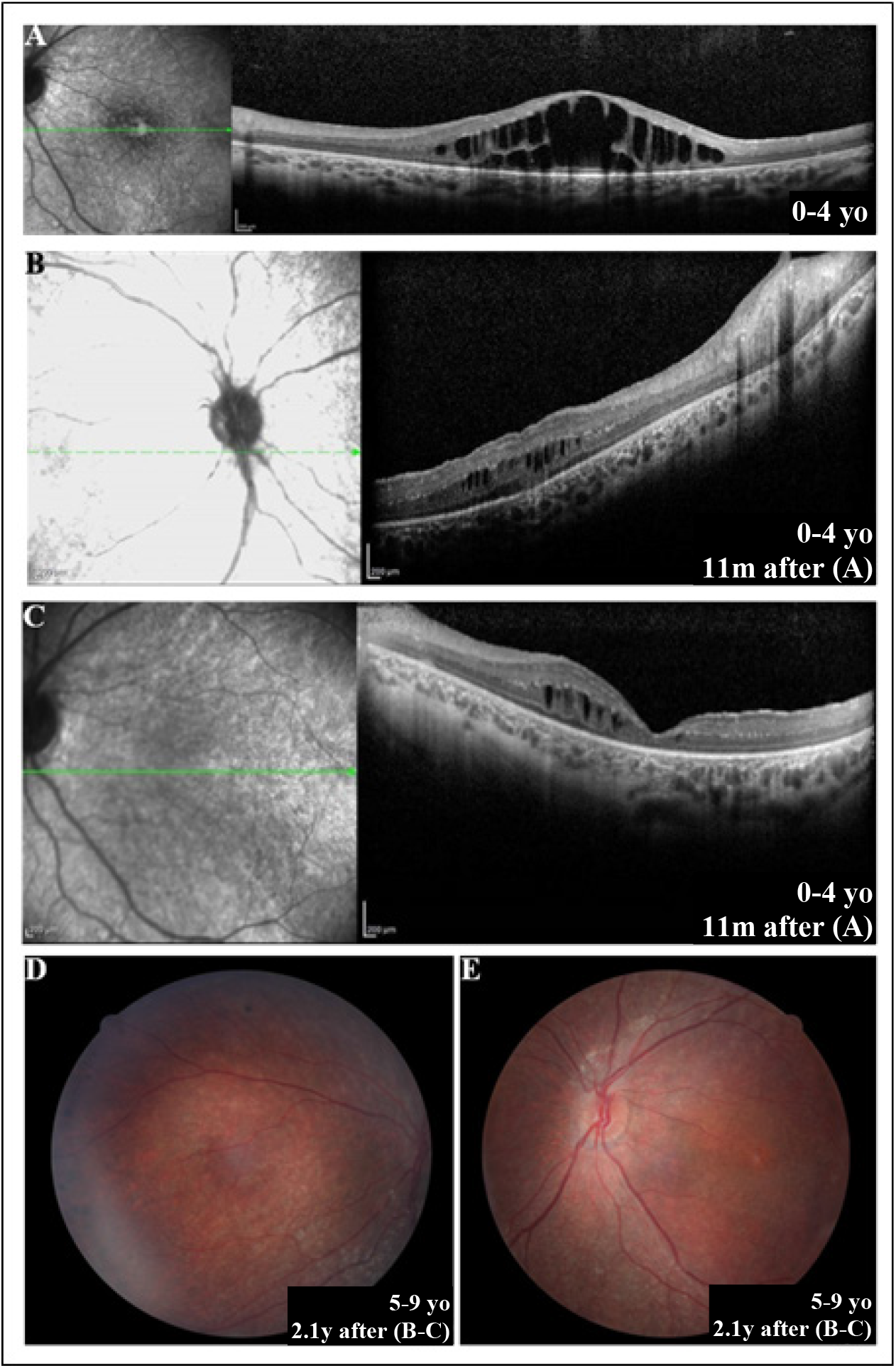
Clinical imaging of Patient 3 from our retrospective cohort with a homozygous c.2528G>A mutation in *PEX1* (p.G843D). A. Oculus dexter (OD) foveal spectral-domain optical coherence tomography (SD-OCT) at age range 0-4 years old (yo) showing marked intra-retinal schitic spaces in both the outer nuclear layer (ONL) and inner nuclear layer (INL). The visual acuity (VA) was 0.75 LogMAR. The schitic spaces in the ONL and INL coalesce centrally. There is otherwise diffuse retinal thinning outside the schitic area. B. OD SD-OCT 11 months after SD-OCT shown in (A) (age range 0-4 yo) (VA data not available) showing marked central foveal thinning as well as shallow schisis in the ONL and small schitic spaces in the INL temporal to the fovea. C. Oculus sinister (OS) SD-OCT 11 months after SD-OCT shown in (A) (age range 0-4 yo) (VA data not available) showing diffuse thinning of the ONL with schitic spaces in the INL temporal to the fovea. D-E. OD and OS color photos 2.1 years after SD-OCT shown in (B, C) (age range 5-9 yo) (VA of 1.18 LogMAR) of the posterior pole showing parafoveal retinal pigment epithelium (RPE) depigmentation, diffuse yellow-beige nummular, leopard-like, RPE pigmentation throughout the posterior pole as well as arteriolar attenuation. Bone-spicules-like-pigmentation can be seen outside the arcades in OD (D). y:years;m:months.

**Figure 5.**
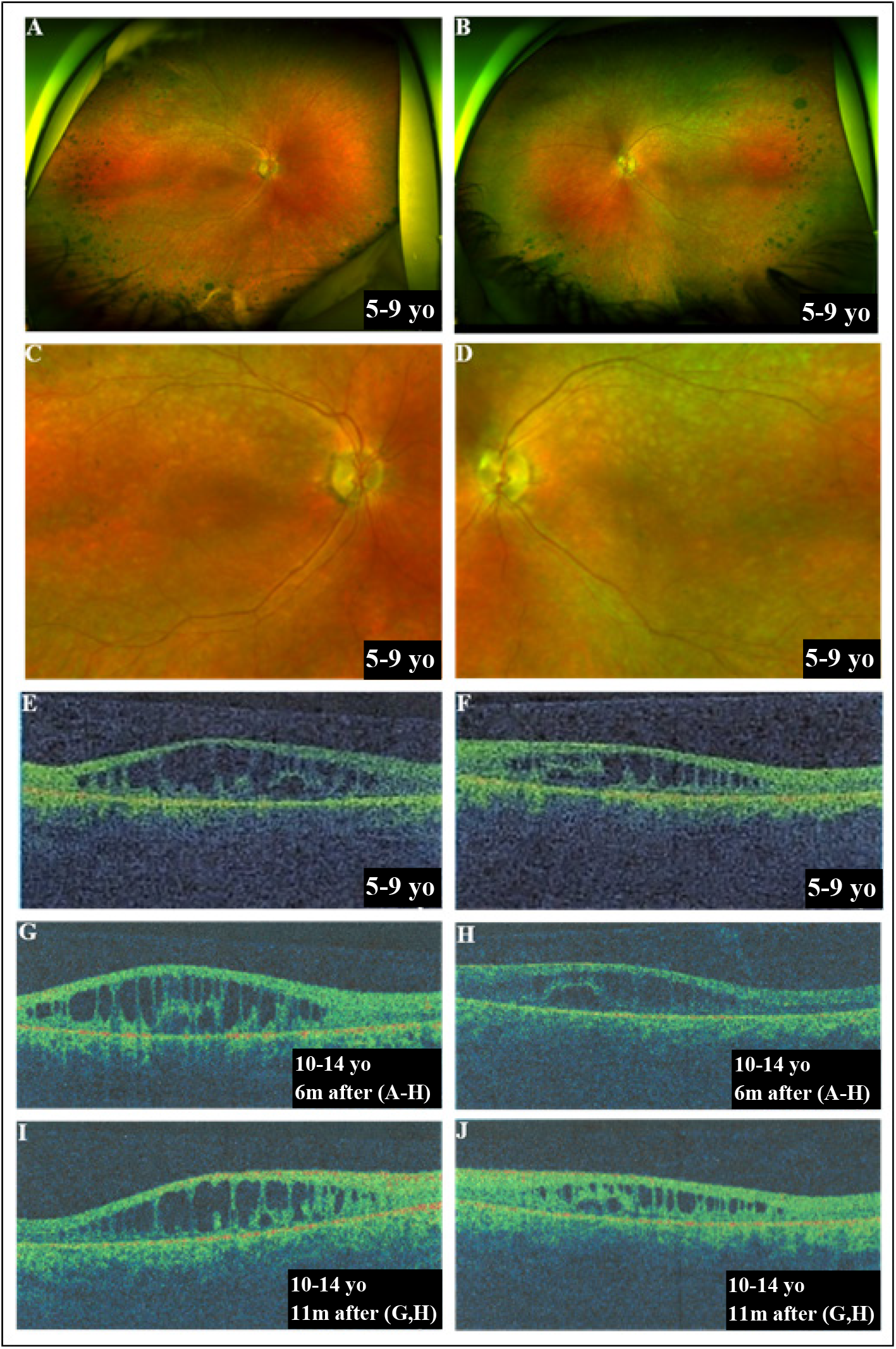
Clinical imagining of Patient 9 from our retrospective cohort with a homozygous c.2528G>A mutation in *PEX1* (p.G843D). A-B. Oculus dexter (OD) and oculus sinister (OS) wide-field color photos at age range 5-9 years old (yo) showing round and oval retinal pigment epithelium (RPE) dark pigmentary changes in the temporal periphery as well as arteriolar attenuation. The visual acuity (VA) was 0.6 LogMAR and 0.7 LogMAR in the OD and OS, respectively. C-D. Magnified color photos of A and B showing characteristic diffuse yellow-beige nummular, leopard-like, RPE pigmentation throughout the posterior pole and arteriolar attenuation. E-F. OD and OS foveal time-domain optical coherence tomography (TD-OCT) at the same age (age range 5-9 yo) showing intra-retinal schitic spaces in both the outer nuclear layer (ONL) and inner nuclear layer (INL) that coalesce centrally. G-I. OD foveal TD-OCT 6 months and 1.5 years later, respectively (age range 10-14 yo), showing the progression and the enlargement of the schitic cavities overtime. H-J. OS foveal TD-OCT at the same age as (G) and (I), respectively (age range 10-14 yo), showing the progression of the schitic cavities overtime. The VA was 0.8 LogMAR in each eye at the age of TD-OCTs shown in (I,J) (age range 10-14 yo). m:months.

**Figure 6.**
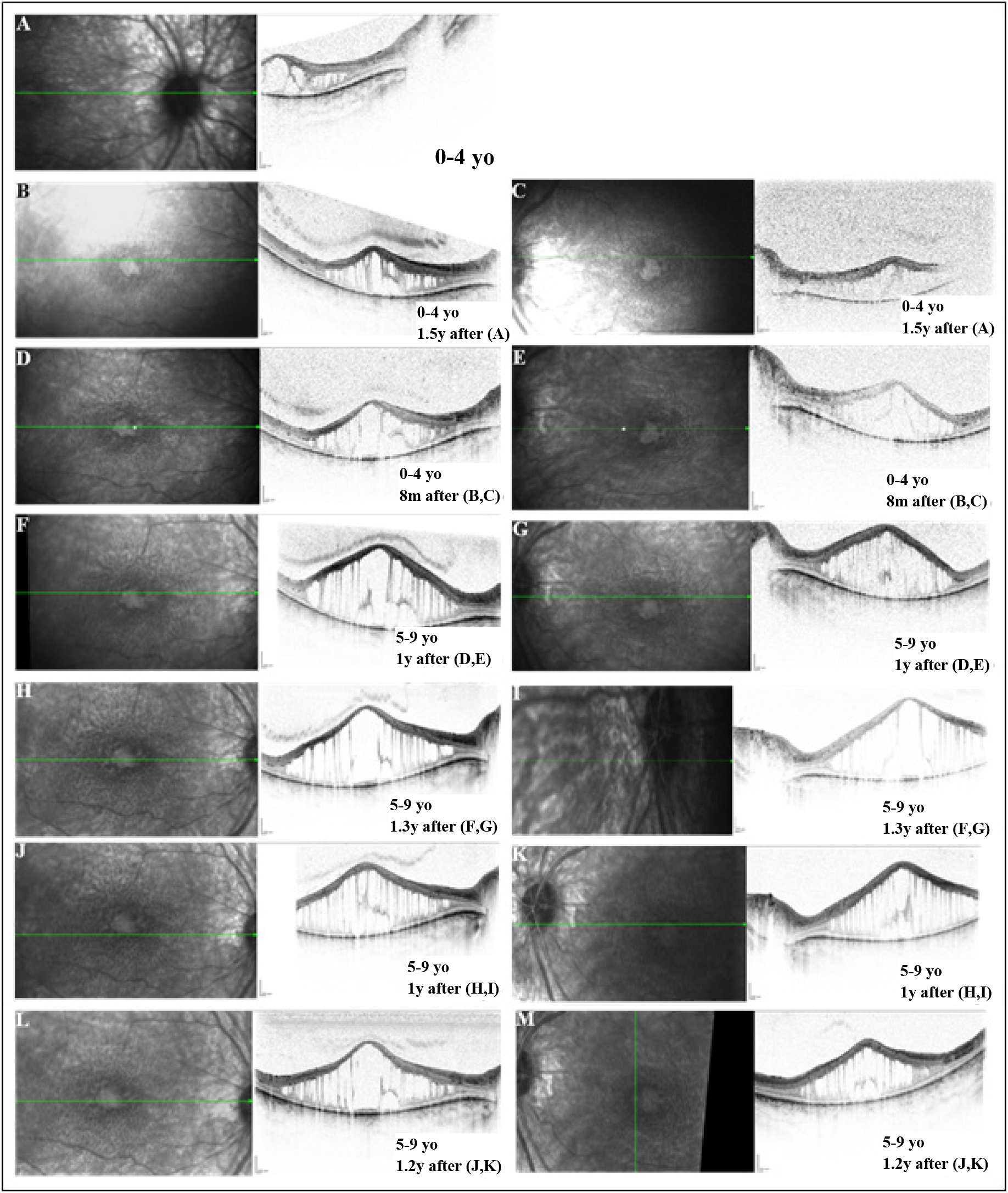
Clinical imagining of Patient 5 from our retrospective cohort with a homozygous c.2528G>A mutation in *PEX1* (p.G843D). A. Oculus dexter (OD) foveal spectral-domain optical coherence tomography (SD-OCT) at age range 0-4 years old (yo) showing marked intra-retinal schitic spaces in both the outer nuclear layer (ONL) and inner nuclear layer (INL). The schitic spaces in the ONL and INL coalesce centrally. There is otherwise diffuse retinal thinning outside the schitic area. B, D, F, H, J and L. OD SD-OCT at subsequent visits showing the progression and the enlargement of the schitic cavities overtime. C, E, G, I, K, and M. Oculus sinister (OS) SD-OCT at subsequent visits showing the progression and the enlargement of the schitic cavities overtime. The visual acuity decreased from 0.7 LogMAR at age range 0-4 yo to 0.9 LogMAR in 6.7 years. m:months; y:years.

**Figure 7.**
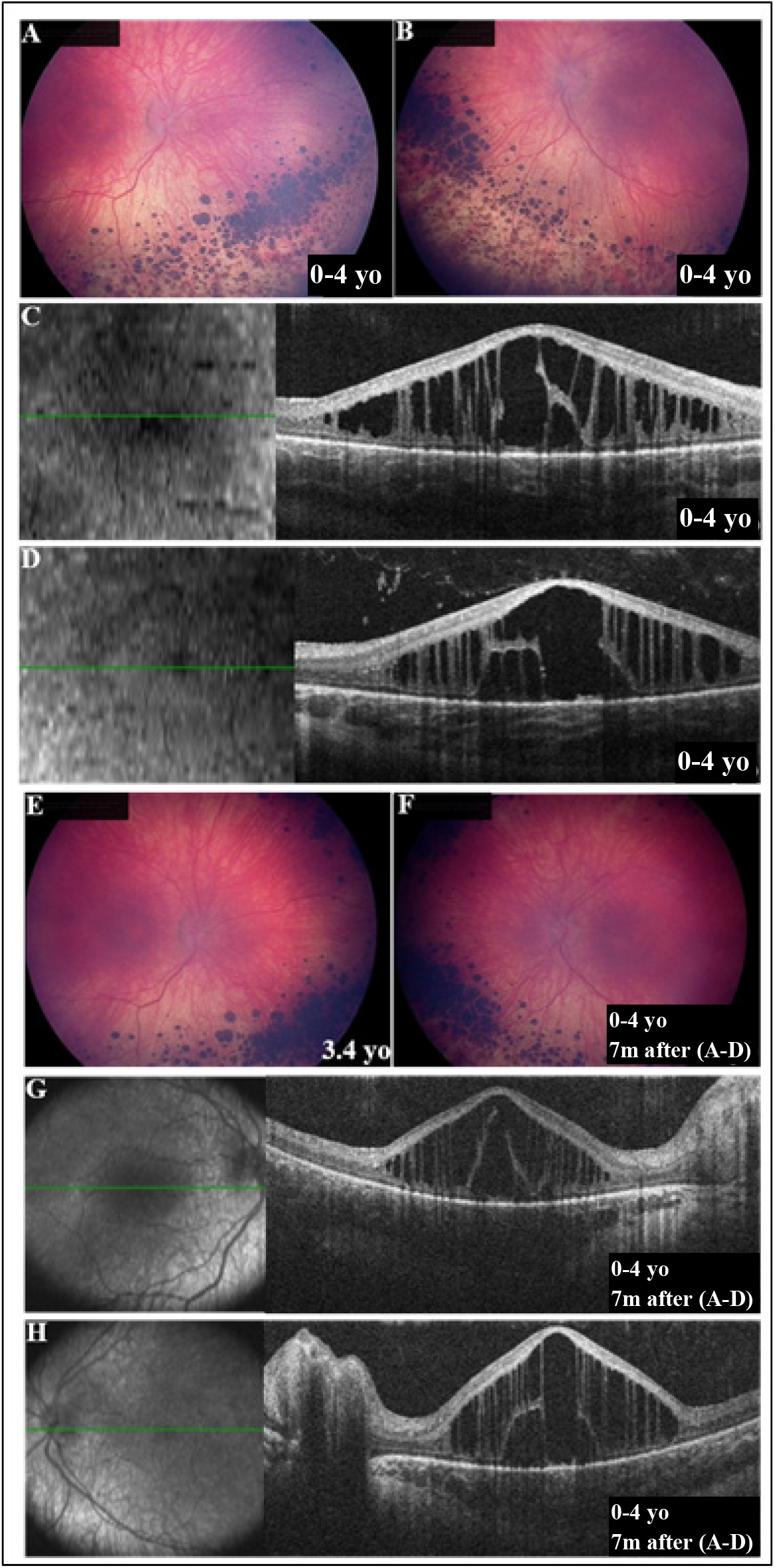
Clinical imagining of Patient 14 from our retrospective cohort with a homozygous c.2528G>A mutation in *PEX1* (p.G843D). A-B. Oculus dexter (OD) and oculus sinister (OS) color photos at age range 0-4 years old (yo) showing round and oval retinal pigment epithelium (RPE) dark pigmentary changes in the infero-nasal quadrant as well as arteriolar attenuation. Both foveas showed RPE depigmentation centrally. The uncorrected visual acuity (VA) was 0.5 LogMAR in each eye. C-D. OD and OS foveal spectral-domain optical coherence tomography (SD-OCT) at the same age (age range 0-4 yo) showing intra-retinal schitic spaces in both the outer nuclear layer (ONL) and inner nuclear layer (INL) that coalesce centrally. E-F. OD and OS color photos 7 months later (age range 0-4 yo) showing interval increase in the number of coalescence of the infero-nasal round and oval RPE dark pigmentation. The uncorrected visual acuity (VA) was 0.9 LogMAR in each eye. G-H. OD and OS foveal SD-OCT at the same age as (E,F) (age range 0-4 yo) showing the progression of intra-retinal ONL and INL schitic spaces. m:months.

**Figure 8.**
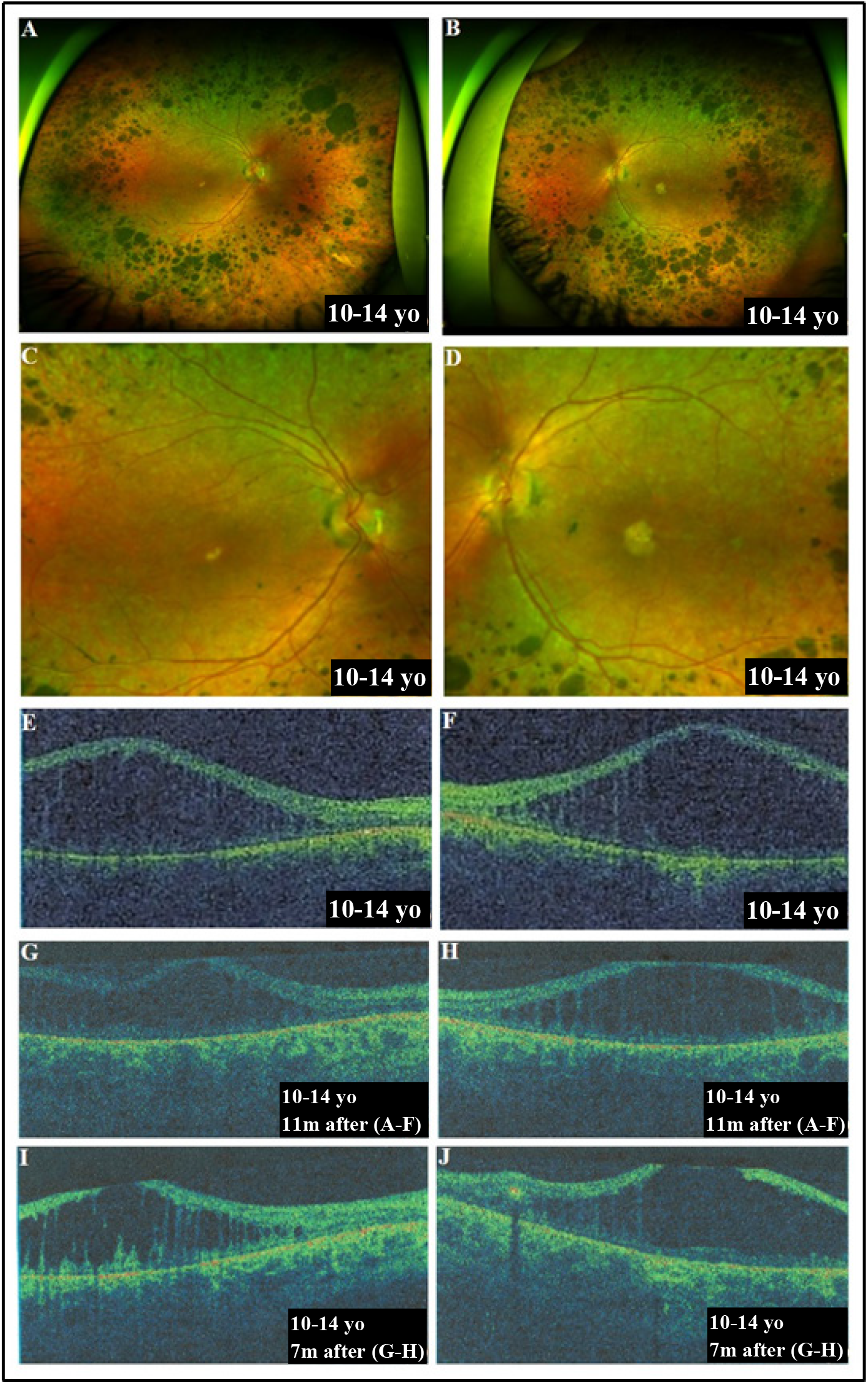
Clinical imagining of Patient 10 from our retrospective cohort with a homozygous c.2528G>A mutation in *PEX1* (p.G843D). A-B. Oculus dexter (OD) and oculus sinister (OS) wide-field color photos at age range 10-14 years old (yo) showing round and oval retinal pigment epithelium (RPE) dark pigmentary changes in a circinate peripheral pattern abutting the arcades. Both fovea showed areas of atrophy. The visual acuity (VA) was 0.8 LogMAR in each eye. C-D. Magnified color photos of A and B showing characteristic diffuse yellow-beige nummular, leopard-like, RPE pigmentation throughout the posterior pole, round and oval RPE dark pigmentation and arteriolar attenuation. E-F. OD and OS foveal time-domain optical coherence tomography (TD-OCT) at the same age (age range 10-14 yo) showing intra-retinal schitic spaces in both the outer nuclear layer (ONL) and inner nuclear layer (INL) that coalesce centrally. G-I. OD foveal TD-OCT 11 months and 1.6 years later, respectively (age range 10-14 yo), showing the progression of the schitic cavities overtime. H-J. OS foveal TD-OCT at the same age as (G) and (I), respectively (age range 10-14 yo), showing the progression of the schitic cavities overtime. The VA was 0.8 LogMAR at the same age as in (I,J) (age range 10-14 yo). m:months.

**Figure 9.**
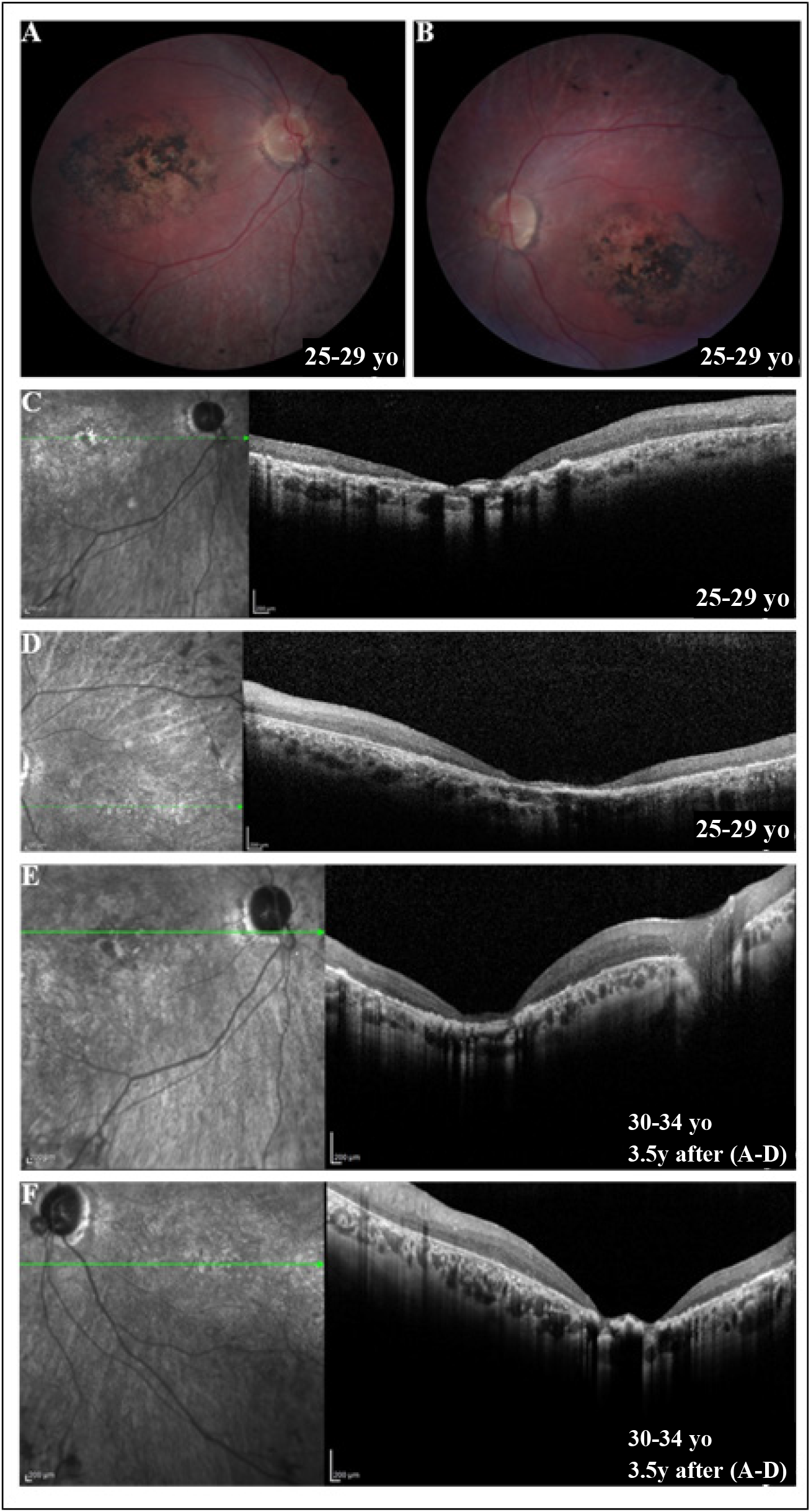
Clinical imagining of Patient 8 from our retrospective cohort with a homozygous c.2528G>A mutation in *PEX1* (p.G843D). A-B. Oculus dexter (OD) and oculus sinister (OS) color photos at age range 25-29 years old (yo) showing central diffuse peri-foveal retinal pigment epithelium (RPE) hyper-pigmentation with atrophy. The uncorrected visual acuity (VA) was 1.15 LogMAR in each eye. C-D. OD and OS foveal spectral-domain optical coherence tomography (SD-OCT) at the same age (age range 25-29 yo) showing marked central peri-foveal atrophy surrounded by diffuse retinal thinning. E-F. OD and OS foveal SD-OCT 3.5 years later (age range 30-34 yo) showing interval mild enlargement of the central peri-foveal atrophy with stable adjacent diffuse retinal thinning. The VA was 3 LogMAR OU. y:years.

## Discussion

In this study, we have presented the common ophthalmological findings reported in 79 patients from the literature and 66 enrolled in our longitudinal natural history study. This represents the largest study of ophthalmological findings in ZSD to date. Furthermore, we stratified these patients into the major ZSD clinical severity groups and thus associated the vision findings with disease severity. Overall, there were fewer examinations reported for severe patients, than intermediate and milder. This could be due to the fact the patients with severe disease generally do not survive infancy.

We determined prevalence in this population for visual acuity, retinal and anterior segment abnormalities, presence of nystagmus, electroretinogram and OCT exams findings. We examined VA across ages using all data, but also per individual over time using our longitudinal data for both VA and retinal structure. Taken together, the data collected indicate that retinal pathology occurs in almost every patient and is the predominant eye pathology in ZSD. Most patients, in which there was data available for these studies, had abnormal pigment spots in the peripheral retina, nystagmus, reduced or extinguished full-field ERGs and intraretinal cysts. We found higher prevalence for these findings in the milder ZSD groups, which could be secondary to these individuals being easier to examine due to older ages and better cognitive abilities, or alternatively, a progressive increase of these findings over time.

Looking at VA over time, we found that around 40% of patients were legally blind, and the median age to blindness was 3.8 years in the intermediate and 7.3 years in the mild group. Linear regression for the logMAR VA vs. age for all patients was statistically significant and showed a slow but progressive decline with age. Intraretinal cysts evolve over time or remain stable. The only patient without intraretinal cyst had her first OCT exam at 28 years and this showed complete loss of the photoreceptor layer^12^.

ZSD is a disorder of global peroxisome dysfunction and peroxisomes are present in all mammalian cells and tissues. Ultrastructure of normal human retina has shown enriched localization of peroxisomes at the base of the photoreceptor outer segment^10^, corresponding to the location of peroxisome enrichment in mouse retinal histology^25,26^. Recently, we (CA, NEB) characterized the murine retina in a knock-in model with the murine equivalent of the common PEX1 p.G843D allele (PEX1 p.G844D). We reported that retinopathy in these mice was marked by an early attenuated cone function and abnormal cone morphology, with subsequent decreasing rod function. Structural defects at the inner retina occurred later in the form of bipolar cell degradation. VA was diminished. This supports that the primary retinal degeneration involves the photoreceptor cells but could be more extensive.

To precisely define the human retinal pathology over time, a systematic, prospective study on vision loss in ZSD is required. This study could also be designed to identify the best clinical endpoints and patient cohorts for use in future interventional trials. This retrospective analysis indicates that full-field ERG may not be a useful measure of visual function in this population since it is uniformly diminished or extinguished (for a single patient over time or among patients), even when patients retain functional vision. More useful measurements could include OCT to monitor retinal structural changes and functional vision measures such as multi-luminance mobility tests and functional vision questionnaires.

Limitations of this study center on the collection of retrospective data from different centers as part of clinical care. Thus, data availability, data type, tests performed, time points and physicians were different amongst patients. Due to the anonymous nature of patient data publication, we cannot preclude that patients were reported more than once, which could lead to over-representation of certain features.

## Supplementary Materials

Supplementary Table 1: Summary of ophthalmic findings in ZSD patients enrolled in our Natural History Study; Supplementary Table 2: Summary of ophthalmic findings in previously reported ZSD patients; Supplementary Table 3: Ophthalmic findings in ZSD patients enrolled in our Natural History Study; Supplementary Table 4: Ophthalmic findings in previously reported Zellweger Spectrum Disorder patients; Supplementary Table 5: List of Zellweger Spectrum Disorder published case reports and cohort studies and their reported ophthalmic findings.

## Data Availability

All data produced in the present study are available upon reasonable request to the authors.

## Acknowledgments

The authors would like to thank the families of the natural history study participants involved in this study for their collaboration and support for research to improve patient care.

## Ethics declaration

The Review Ethics Board of the McGill University Health Center gave ethical approval for this work (Study #11-090-PED). Participants or their legal representatives provided their consent to be part of the Natural History study #11-090-PED and to have the results of this study published. Participants or their legal representative provided their authorizations for obtaining their medical records for this study.

## Conflict of Interest Disclosures

No authors have relevant financial disclosures involving the work under consideration for publication.

### Author Contributions

All authors (CY, RGC, FA, CA, RKK and NEB) all had full access to all the data in the study and take responsibility for the integrity of the data and the accuracy of the data analysis. Study concept and design, CY, RGC, CA, RKK and NEB; Acquisition of data (Scoping literature review), RGC, FA, CY, CA; Acquisition of data (Retrospective medical chart review), CY; Analysis and interpretation of data: CY, RGC, FA, RKK and NEB; Drafting of the manuscript: CY, RGC, FA, CA and NEB; Critical revision of the manuscript for important intellectual content: CY, RGC, FA, CA, RKK and NEB, RKK; Administrative, technical, and material support: NEB.

