## Supplementary Table 1 for "Peroxisome Biogenesis Disorders in the Zellweger Spectrum: Ophthalmic Findings from a New Natural History Study Cohort and Scoping Literature Review"

**Supplementary Table 1. Summary of ophthalmic findings in ZSD patients enrolled in our Natural History Study.** Retrospective data collected from medical records of 66 patients. The proportion of patients with a specific ophthalmic finding out of number of patients with assessment is shown, with median ages at finding shown in parentheses. Abbreviations: ERG = Electroretinogram; OCT = Optical Coherence Tomography; VA = Visual Acuity.
*One of the following : microspherophakia, white deposits in vitreous humor, corneal opacities, keratopathy, corneal abrasion, uveitis, dry cornea, asteroid hyalosis, aphakia, lens subluxation, lens opacities.

| | | Proportion of patients with assessment (median age, y) | | | |
|---|---|---|---|---|---|
| **Ocular findings – Natural History Study** | **Total patients** | **Severe ZSD** | **PEX1 p.G843D/Null - Intermediate ZSD** | **Mild-Intermediate ZSD** | **PEX1 G843D/G843D - Mild ZSD** |
| **Total patients** | **66** | **4** | **20** | **27** | **15** |
| Mean age at first ophthalmological exam | **3** | 0.3 | 3.8 | 2.6 | 3.6 |
| **Visual acuity (VA) – Number of patients** | **20** | 0 | **3** | **5** | **12** |
| Number of patients with VA < 1 (Median age at onset) | 16/20 (8.3) | 0 | 2/3 (4.4) | 4/5 (7.5) | 10/12 (8.9) |
| Number of patients with legal blindness | 10/20 | 0 | 2/3 | 1/5 | 7/12 |
| Mean age of onset of legal blindness (y) | 8.4 | N/A | 3.8 | 10.8 | 9.4 |
| **Anterior segment – Number of patients** | **43** | **4** | **10** | **20** | **9** |
| Normal | 27/43 | 1/4 (0.4) | 5/10 (5.2) | 15/20 (2.8) | 6/9 (4.3) |
| Cataracts | 7/43 | 2/4 (0.3) | 2/10 (22.4) | 1/20 (10.8) | 2/9 (18.7) |
| Other anterior segment abnormalities* | 11/43 | 1/4 (0.1) | 4/10 (19.3) | 4/20 (6.1) | 2/9 (27.5) |
| Not assessed | 23/66 | 1/5 | 9/19 | 7/27 | 6/15 |
| **Fundus – Number of patients** | **59** | **3** | **16** | **25** | **15** |
| Normal | 7/59 | 0 | 0/16 | 7/25 (3.6) | 0/15 |
| Abnormal pigmentation in periphery | 38/59 | 2/3 (0.7) | 11/16 (9.2) | 10/25 (4.3) | 15/15 (7.4) |
| Optic disc pallor | 27/59 | 1/3 (0.2) | 10/16 (14.2) | 6/25 (3.1) | 10/15 (13) |
| Maculopathy | 20/59 | 1/3 (0.2) | 3/16 (4.9) | 5/25 (3.2) | 11/15 (10.3) |
| Attenuated retinal vessels | 19/59 | 1/3 (0.4) | 7/16 (8) | 3/25 (6.2) | 8/15 (17.4) |
| Retinal degeneration | 21/59 | 0 | 6/16 (4.3) | 7/25 (4) | 8/15 (7) |
| Not assessed | 7/66 | 2/5 | 3/19 | 2/27 | 0/15 |
| **Nystagmus – Number of patients** | **58** | **4** | **17** | **22** | **15** |
| Yes | 44/58 | 1/4 (0.3) | 15/17 (1.7) | 16/22 (3.3) | 12/15 (4.4) |
| No | 14/58 | 3/4 (0.2) | 2/17 (3.9) | 6/22 (2.7) | 3/15 (26.3) |
| Not assessed | 8/66 | 1/5 | 2/19 | 5/27 | 0/15 |

Supplementary Table 1.

| | | | | | |
|---|---|---|---|---|---|
| **Refraction – Number of patients** | **48** | **0** | **16** | **18** | **14** |
| Number of patients with hyperopia | 38/48 | 0 | 16/16 | 8/18 | 14/14 |
| Mean spherical refractive error | 4.10 | N/A | 3.15 | 4.77 | 4.21 |
| **Number of ERGs (Number of patients)** | **16 (8)** | **0** | **0** | **4 (1)** | **12 (7)** |
| Flat/Not detectable (mean age at onset, y) | 9 (5.2) | 0 | 0 | 3 (4.3) | 6 (6.5) |
| Reduced photopic and scotopic responses (mean age at onset, y) | 7 (5.5) | 0 | 0 | 1 (6) | 6 (6.6) |
| Delayed peak times | 1 | 0 | 0 | 0 | 1 |
| **Number of OCTs (Number of patients)** | **28 (8)** | **0** | **0** | **6 (2)** | **22 (6)** |
| Schitic changes (mean age at onset, y) | 21 (6.9) | 0 | 0 | 6 (7.4) | 15 (9.2) |
| Absence of photoreceptor layer (mean age at onset, y) | 2 (1.8) | 0 | 0 | 0 | 2 (1.8) |
