## Supplementary Table 2 for "Peroxisome Biogenesis Disorders in the Zellweger Spectrum: Ophthalmic Findings from a New Natural History Study Cohort and Scoping Literature Review"

**Supplementary Table 2. Summary of ophthalmic findings in previously reported ZSD patients.** The proportion of patients with a specific ophthalmic finding out of number of patients with assessment is shown, with age ranges reported shown in parentheses. Abbreviations: F+F = Fix and Follow; LP = Light perception; PR = Photoreceptors; VA = Visual Acuity.

| | Proportion of patients with assessment (age range, y) | | | | |
|---|---|---|---|---|---|
| **Ocular findings** | **Total # of patients** | Severe ZSD | PEX1 p.G843D/Null - Intermediate ZSD | Mild-Intermediate ZSD | PEX1 p.G843D/G843D - Mild ZSD |
| **Total patients** | **79** | **13** | **2** | **50** | **14** |
| **Visual acuity** | **48** | 2 | 2 | 32 | 12 |
| Normal | **1/48** | 0/2 | 0/2 | 1/32 (7) | 0/12 |
| LP | **4/48** | 1/2 (0.06) | 0/2 | 3/32 | 0/12 |
| F+F | **9/48** | 1/2 (0.6) | 1/2 (2) | 7/32 (0.9 – 9) | 0/12 |
| < 1.00 | **17/48** | 0/2 | 0/2 | 11/32 (4 – 30) | 6/12 (8 – 35) |
| ≥ 1.00 | **17/48** | 0/2 | 1/2 (18) | 10/32 (7 – 27.5) | 6/12 (22 – 32) |
| Not assessed | **31/79** | 11/13 | 0/2 | 18/50 | 2/14 |
| **Anterior segment** | **21** | 8 | 0 | 12 | 1 |
| Normal | **5/21** | 1/8 (0.06) | 0 | 4/12 (0.6 – 13) | 0/1 |
| Cataracts | **12/21** | 5/8 (4 do – 0.8) | 0 | 6/12 (1.7 – 27.5) | 1/1 (25) |
| Cloudy cornea | **3/21** | 2/8 (0.6 – 1.2) | 0 | 1/12 (4) | 0/1 |
| Not assessed | **58/79** | 5/13 | 2/2 | 38/50 | 13/14 |
| **Fundus** | **71** | 12 | 2 | 45 | 12 |
| Normal | **3/71** | 1/12 (0.6) | 0/2 | 2/45 (6) | 0/12 |
| Abnormal pigmentation in periphery | **58/71** | 9/12 (0.06 - 1.5) | 2/2 (2 – 1.3) | 35/45 (0.25 – 30) | 12/12 (8 – 35) |
| Optic disc pallor | **9/71** | 2/12 (0.5 – 1.5) | 0/2 | 6/45 (1.5 – 30) | 1/12 (1.3) |
| Maculopathy | **18/71** | 5/12 (4 do – 1.2) | 0/2 | 12/45 (0.6 – 21) | 1/12 (1.3) |
| Attenuated vessels | **15/71** | 2/12 (0.06 – 0.5) | 0/2 | 11/45 (0.4 – 30) | 2/12 (0.7 – 1.3) |
| Retinal degeneration | **15/71** | 3/12 (4 do – 1.2) | 0/2 | 11/45 (1.7 – 12) | 1/12 (0.7) |
| Not assessed | **8/79** | 1/13 | 0/2 | 5/50 | 2/14 |
| **Nystagmus** | **37** | 3 | 2 | 28 | 4 |
| No | **16/37** | 0/3 | 0/2 | 16/28 (0.9 – 15) | 0/4 |
| Yes | **21/37** | 3/3 (0.06) | 2/2 (2 – 18) | 12/28 (0.4 – 19.5) | 4/4 (1.3 - 32) |
| Not assessed | **42/79** | 10/13 | 0/2 | 22/50 | 10/14 |
| **ERG** | **48** | 10 | 0 | 35 | 3 |
| Normal amplitude | **2/48** | 0/10 | 0 | 2/35 (17 – 29) | 0/3 |
| Attenuated amplitude | **25/48** | 1/10 (0.03) | 0 | 23/35 (0.4 – 30) | 1/3 (25) |

Supplementary Table 2.

| | | | | | | |
|---|---|---|---|---|---|---|
| | Extinguished | **18/48** | 9/10 (0.19 – 1.5) | 0 | 7/35 (0.6 – 21) | 2/3 (8 – 28) |
| | Normal latency | **2/48** | 0/10 | 0 | 2/35 (1.5 – 9) | 0/3 |
| | Delayed latency | **7/48** | 0/10 | 0 | 7/35 (0.9 - 30) | 0/3 |
| | Not assessed | **31/79** | 3/13 | 2/2 | 15/50 | 11/14 |
| **VEP** | | **28** | 7 | 0 | 21 | 0 |
| | Normal P-100 amplitude | **12/28** | 0/7 | 0 | 12/21 (0.9 – 13) | 0 |
| | Attenuated P-100 amplitude | **12/28** | 5/7 (0.06 – 1.2) | 0 | 7/21 (0.25 – 7) | 0 |
| | Normal P-100 latency | **9/28** | 0/7 | 0 | 9/21 (0.9 – 9) | 0 |
| | Delayed P-100 latency | **9/28** | 3/7 (0.06 - 1.5) | 0 | 6/21 (1.5 – 4.3) | 0 |
| | Not assessed | **51/79** | 6/13 | 2/2 | 29/50 | 14/14 |
| **OCT** | | **13** | 0 | 0 | 9 | 4 |
| | No intraretinal cysts | **4/13** | 0 | 0 | 3/9 (1.7 – 12) | 1/4 (28) |
| | Intraretinal cysts | **8/13** | 0 | 0 | 5/9 (6 – 30) | 3/4 (4 – 25) |
| | Depletion of PRs | **6/13** | 0 | 0 | 4/9 (1.7 – 12) | 2/4 (8 - 28) |
| | Not assessed | **66/79** | 13/13 | 2/2 | 41/50 | 10/14 |
