## Supplementary Table 3 for "Peroxisome Biogenesis Disorders in the Zellweger Spectrum: Ophthalmic Findings from a New Natural History Study Cohort and Scoping Literature Review"

**Supplementary Table 3. Ophthalmic findings in ZSD patients enrolled in our Natural History Study.** Retrospective data collected from medical records of 66 patients and shown in chronological order for each patient. Empty cells represent no available data. Presence of the finding is represented by "+" and absence of the finding is represented by "-". Grey sections show the variation of VA (ΔVA) between the first and last assessment for the patient (ΔT) and shows the calculated variation of VA per year (ΔVA/year). Fundus and OCT descriptions are reported as written in the patient's medical charts. Abbreviations: Abn = Abnormal; Avg = Average; CAI = Carbonic Anhydrase Inhibitor; CME = cystoid macular edema; INL = inner nuclear layer; OD = right eye; ONL = outer nuclear layer; OS: left eye; OU = both eyes; VA = Visual Acuity

| Patient # | Disease severity subgroup | Mutations | Sex | Age range (time since previous records) (y) | Avg VA ALL (LogMar) | Anterior segment | Fundus | | | | | | Nystagmus | OCT Description |
|---|---|---|---|---|---|---|---|---|---|---|---|---|---|---|
| | | | | | | | Normal | Abn.pigmentation in peripheral retina | Optic disc pallor | Attenuated vessels | Retinal degeneration | Fundus – Other descriptions | | |
| 1 | PEX1 p.G843D / G843D - Mild ZSD | PEX1 c.2528G>A (p.G843D) / c.2528G>A (p.G843D) | F | 5-9 | 0.3 | | | + | | | | Macular dystrophy, Optic nerve atrophy | | |
| | | | | 5-9 (1.8) | 0.575 | | | | | | | | | |
| | | | | 5-9 (0.2) | | | | + | | | | Maculopathy | + | |
| | | | | 5-9 (0.3) | | Normal | | | | | | | | |
| | | | | 5-9 (0) | 0.725 | | | | | | | | | |
| | | | | 10-14 (0.6) | 0.75 | Normal | | + | | | | Macular pigmentary changes | | |
| | | | | 10-14 (0.2) | 0.75 | | | | | | | | | |
| | | | | 10-14 (0.8) | 0.775 | Normal | | + | | | | Macular pigmentary changes | | |
| | | *ΔT (y)* | | *3.9* | | | | | | | | | | |
| | | *ΔVA* | | | *0.475* | | | | | | | | | |
| | | *ΔVA/year* | | | *0.122* | | | | | | | | | |
| 2 | PEX1 p.G843D / G843D - Mild ZSD | PEX1 c.2528G>A (p.G843D) / c.2528G>A (p.G843D) | M | 0-4 | | | | + | + | + | | | + | |
| (Patient 1's sibling) | | | | 0-4 (0.8) | | | | | | | | | + | |
| | | | | 0-4 (0.2) | | | | + | + | + | | Maculopathy | + | |
| | | | | 0-4 (3.5) | | | | | | | | | + | |
| 3 | PEX1 p.G843D / G843D - Mild ZSD | PEX1 c.2528G>A (p.G843D) / c.2528G>A (p.G843D) | M | 0-4 | | Normal | | + | | | | | + | |
| | | | | 0-4 (0.3) | | | | | | | | | + | |
| | | | | 0-4 (2.9) | 0.95 | | | + | | | | Maculopathy | | |
| | | | | 5-9 (2.9) | 0.95 | | | | | | + | Macular edema | | Advanced CME |
| | | | | 5-9 (1.3) | | | | | | | | Maculopathy | + | |
| | | | | 5-9 (0.1) | | | | | | | | Macular edema | | Advanced CME |
| | | *ΔT (y)* | | *2.9* | | | | | | | | | | |
| | | *ΔVA* | | | *0* | | | | | | | | | |
| | | *ΔVA/year* | | | *0* | | | | | | | | | |
| 4 | PEX1 p.G843D / G843D - Mild ZSD | PEX1 c.2528G>A (p.G843D) / c.2528G>A (p.G843D) | F | 0-4 | | | | + | | | | | + | |
| | | | | 0-4 (0.5) | | | | | | | | | + | |
| | | | | 0-4 (1.7) | | | | | | | | | - | |
| | | | | 15-19 (13.2) | 1 | | | + | | | | | | |
| | | | | 25-29 (8.7) | | | | + | | | + | | - | |
| | | | | 25-29 (0.1) | | | | | | | | | - | Macular edema OU |

Supplementary Table 3.

| Patient # | Disease severity subgroup | Mutations | Sex | Age range (time since previous records) (y) | Avg VA ALL (LogMar) | Anterior segment | Fundus | | | | | | Nystagmus | OCT Description |
|---|---|---|---|---|---|---|---|---|---|---|---|---|---|---|
| | | | | | | | Normal | Abn.pigmentation in peripheral retina | Optic disc pallor | Attenuated vessels | Retinal degeneration | Fundus – Other descriptions | | |
| | | | | 25-29 (0.3) | | | | | | | | Maculopathy | - | |
| | | | | 25-29 (0.2) | | | | | | | | Macular edema | - | |
| | | | | 25-29 (0.5) | | | | | | | | | - | |
| | | | | 25-29 (0.6) | | | | | | | | Macular edema | - | |
| | | | | 25-29 (0.4) | | | | | | | | Macular edema, Dry macula | - | |
| | | | | 25-29 (0.3) | | Normal | | | | | | Less papillomacular bundle | - | |
| | | | | 25-29 (0.6) | | Cataracts | | | | | | | | |
| | | | | 25-29 (0) | | Early posterior subcapsular cataracts | | | | | | Intraretinal edema, Optic nerve drusen | - | |
| | | | | 25-29 (0.4) | | | | | | | | | - | |
| 5 | PEX1 p.G843D / G843D - Mild ZSD | PEX1 c.2528G>A (p.G843D) / c.2528G>A (p.G843D) | M | 0-4 | | Normal | | + | | | + | Blond periphery, Small optic nerve | - | |
| | | | | 0-4 (0.2) | | | | | | | | | | Extensive swelling in the fovea (OD) and absence of photoreceptor layer (OU). Marked intra-retinal schitic spaces in both the outer nuclear layer (ONL) and inner nuclear layer (INL). The schitic spaces in the ONL and INL coalesce centrally. There is otherwise diffuse retinal thinning outside the schitic area. |
| | | | | 0-4 (0.4) | | Normal | | + | + | | | Small optic nerve | | |
| | | | | 0-4 (0.4) | | Normal | | + | + | | | Small optic nerve | | |
| | | | | 0-4 (0.1) | | | | + | + | | | Blond periphery, Small optic nerve | | |
| | | | | 0-4 (0.3) | | | | | | | | | | CME and absence of photoreceptor layer. Starting CAI eye drops (Dorzolamide) |
| | | | | 0-4 (0.1) | 0.6 | Normal | | + | + | | | Blond periphery, Small optic nerve | | |
| | | | | 0-4 (0.2) | 0.7 | Normal | | | | | | | | Continued edema. Swelling of fovea OU |
| | | | | 0-4 (0.1) | | Normal | | + | + | | | Optic nerve atrophy, Blond periphery | | CME. Stopped CAI eye drops (Dorzolamide). |
| | | | | 0-4 (0) | | | | | + | | | | | |
| | | | | 0-4 (0.3) | 2 | Normal | | + | + | | | Optic nerve atrophy, Blond periphery | | |
| | | | | 0-4 (0.3) | | | | | | | | | | Stable with continued significant edema and retinoschisis OU. Swelling of fovea OU. |
| | | | | 0-4 (0.1) | 1 | Normal | | + | + | | | Optic nerve atrophy, Blond periphery | | |
| | | | | 0-4 (0.5) | | | | + | + | | | Blond periphery, Small optic nerve | | |
| | | | | 0-4 (0.1) | 1 | | | | | | | | | |
| | | | | 5-9 (0.3) | 1.3 | Normal | | + | | | | Optic nerve atrophy, Edema, Blond periphery | | Schisis with edema OD > OS |
| | | | | 5-9 (0.2) | 1.2 | | | | | | | | | |
| | | | | 5-9 (0.4) | 1.25 | | | | | | | | | |
| | | | | 5-9 (0) | 0.9 | Normal | | + | | | | Optic nerve atrophy, Blond periphery | | Stable CME. |

Supplementary Table 3.

| Patient # | Disease severity subgroup | Mutations | Sex | Age range (time since previous records) (y) | Avg VA ALL (LogMar) | Anterior segment | Fundus: Normal | Fundus: Abn.pigmentation in peripheral retina | Fundus: Optic disc pallor | Fundus: Attenuated vessels | Fundus: Retinal degeneration | Fundus – Other descriptions | Nystagmus | OCT Description |
|---|---|---|---|---|---|---|---|---|---|---|---|---|---|---|
| | | | | 5-9 (0.4) | 1.15 | Normal | | + | | | | Blond periphery, Small optic nerve | | |
| | | | | 5-9 (0) | 1.05 | | | | | | | | | |
| | | | | 5-9 (0.1) | 1.15 | | | | | | | | | |
| | | | | 5-9 (0.2) | 0.7 | | | | | | | | | |
| | | | | 5-9 (0.2) | 0.7 | Normal | | | | | | Optic nerve atrophy | | |
| | | | | 5-9 (0.4) | 0.7 | | | | | | | | | |
| | | | | 5-9 (0.4) | 0.5 | | | | | | | | | |
| | | | | 5-9 (0.1) | 0.85 | | | + | | | | | + | |
| | | | | 5-9 (0.2) | 0.7 | | | | | | | | | |
| | | | | 5-9 (0.3) | 1.1 | | | + | | | + | | | |
| | | | | 5-9 (0.5) | 0.925 | | | + | | | | Small optic nerve | + | |
| | | | | 5-9 (1) | 1.025 | | | + | | | | Small optic nerve | | |
| | | | *ΔT (y)* | *6.3* | | | | | | | | | | |
| | | | *ΔVA* | | *0.425* | | | | | | | | | |
| | | | *ΔVA/year* | | *0.067* | | | | | | | | | |
| 6 | PEX1 p.G843D / G843D - Mild ZSD | PEX1 c.2528G>A (p.G843D) / c.2528G>A (p.G843D) | F | 0-4 | 0.55 | | | | | | | | + | |
| | | | | 0-4 (0.6) | | | | | | | | | + | |
| | | | | 0-4 (0.2) | | | | | | | | | + | |
| | | | | 0-4 (2.4) | | | | + | + | | | | + | |
| | | | | 5-9 (1) | | | | | | | | | + | |
| | | | | 5-9 (0.1) | | | + | | | | | | | |
| | | | | 5-9 (2.1) | 1.1 | | | | | | | | | |
| | | | *ΔT (y)* | *6.4* | | | | | | | | | | |
| | | | *ΔVA* | | *0.55* | | | | | | | | | |
| | | | *ΔVA/year* | | *0.086* | | | | | | | | | |
| 7 | PEX1 p.G843D / G843D - Mild ZSD | PEX1 c.2528G>A (p.G843D) / c.2528G>A (p.G843D) | M | 5-9 | 0.78 | | | | | | | | | |
| | | | | 5-9 (1.2) | 0.75 | | | | | | | | | |
| | | | | 10-14 (2.1) | 1 | | | | | | | | | |
| | | | | 10-14 (0.4) | 1 | | | + | | | | | | |
| | | | | 10-14 (0.3) | 1 | | | | | | | | | |
| | | | | 10-14 (0.3) | | | | + | | + | | Atrophy of RPE, gliosis, Small optic discs | | |
| | | | | 10-14 (1.1) | 1 | Lens with punctate nuclear changes | | + | + | + | | Macular hyperpigmentation | | |
| | | | | 15-19 (1.6) | 1 | Normal | | | | | | Macular pigmentary changes | | |
| | | | | 15-19 (0.1) | 0.475 | Traces of lens opacities | | + | | | | | | |

Supplementary Table 3.

| Patient # | Disease severity subgroup | Mutations | Sex | Age range (time since previous records) (y) | Avg VA ALL (LogMar) | Anterior segment | Fundus | | | | | | Nystagmus | OCT Description |
|---|---|---|---|---|---|---|---|---|---|---|---|---|---|---|
| | | | | | | | Normal | Abn.pigmentation in peripheral retina | Optic disc pallor | Attenuated vessels | Retinal degeneration | Fundus – Other descriptions | | |
| | | | | 15-19 (1.2) | 1 | | | | | | | | | |
| | | | | 15-19 (0.6) | 1 | Tiny posterior subcapsular cataract. | | + | + | | | | | |
| | | | | 15-19 (0.1) | 0.6 | Posterior subcapsular cataracts with multi white flecks in nucleus and cortex OU | | + | + | + | | Maculopathy | | |
| | | | | 15-19 (0.1) | 1 | Posterior subcapsular cataracts with multi white flecks in nucleus and cortex OU. No lens subluxation. | | | | + | | Optic nerve atrophy | | |
| | | | | 15-19 (1.1) | 1 | Lens subluxation | | + | + | + | | | - | |
| | | | | 15-19 (0.7) | 0.8 | Many punctate white lens opacities OU. | | | | | | | | |
| | | | | 15-19 (0.6) | 0.85 | Lens cataracts OS>OD. Lens subluxation. | | + | + | + | | | | |
| | | | | 20-24 (1.1) | 1 | Lens with cortical flecks OS>OD w subluxation | | + | + | + | | Macular atrophy | + | |
| | | | | 30-34 (11.9) | | | | | | | | | + | |
| | | | | 30-34 (1) | | | | | | | | | - | |
| | | | | 35-39 (5.1) | | Pseudophakia intraocular lens well centered | | | + | | | Optic neuropathy, nonexudative atrophic dry macular degeneration | | |
| | | | | 35-39 (0.5) | | Pseudophakia intraocular lens well centered | | | + | | | Nonexudative-atrophic dry macular degeneration. Glaucoma primary open angle. | | |
| | | | | 35-39 (0.4) | | | | | + | | | | | |
| | | | | 40-44 (0.7) | | Pseudophakia intraocular lens well centered | | | + | | | Nonexudative age-related macular degeneration. Primary open-angle glaucoma, moderate stage. | | |
| | | | | 40-44 (0.5) | | Pseudophakia intraocular lens well centered | | | + | | | Primary open-angle glaucoma, moderate stage. Nonexudative age-related macular degeneration. | | Pale optic nerve |
| | | | *ΔT (y)* | *12.6* | | | | | | | | | | |
| | | | *ΔVA* | | *0.22* | | | | | | | | | |
| | | | *ΔVA/year* | | *0.017* | | | | | | | | | |
| 8 | PEX1 p.G843D / G843D - Mild ZSD | PEX1 c.2528G>A (p.G843D) / c.2528G>A (p.G843D) | F | 0-4 | | | | + | | | | | | |
| | | | | 0-4 (0.3) | | | | + | | | | | | |
| | | | | 0-4 (0.5) | 0.85 | | | | | | | | + | |
| | | | | 0-4 (0.1) | 0.85 | | | + | | + | | Macular pigmentary changes | + | |

Supplementary Table 3.

| **Patient #** | **Disease severity subgroup** | **Mutations** | **Sex** | **Age range (time since previous records) (y)** | **Avg VA ALL (LogMar)** | **Anterior segment** | **Fundus** | | | | | | **Nystagmus** | **OCT Description** |
|---|---|---|---|---|---|---|---|---|---|---|---|---|---|---|
| | | | | | | | Normal | Abn.pigmentation in peripheral retina | Optic disc pallor | Attenuated vessels | Retinal degeneration | Fundus – Other descriptions | | |
| | | | | 0-4 (0.3) | 0.7 | | | | | | | | | |
| | | | | 5-9 (0.8) | 0.65 | | | | | | | | | |
| | | | | 5-9 (0.5) | 0.9 | | | | | | | | | |
| | | | | 5-9 (0.1) | 1.05 | | | | | | | | + | |
| | | | | 5-9 (0.1) | 1 | | | | | | | | + | |
| | | | | 5-9 (0.2) | | | | + | | | | Macular pigmentary changes | | |
| | | | | 5-9 (0.5) | 1 | | | | | | | | | |
| | | | | 5-9 (0.5) | 1 | | | | | | | | | |
| | | | | 5-9 (1) | 0.85 | | | | | | | | | |
| | | | | 5-9 (1.2) | 1 | | | | | | | | | |
| | | | | 10-14 (1) | 0.825 | | | | | | | | + | |
| | | | | 10-14 (1.5) | 1 | | | + | | | | Optic nerve drusen OS | | |
| | | | | 10-14 (1.3) | 0.825 | | | | | | | Optic nerve drusen OS | | |
| | | | | 10-14 (1.1) | 0.9 | | | | | | | | | |
| | | | | 15-19 (1.3) | 0.85 | | | | | | | | + | |
| | | | | 15-19 (0.6) | 1 | | | | | | | | | |
| | | | | 15-19 (0.4) | 0.825 | | | | | | | | | |
| | | | | 15-19 (0.6) | 0.875 | | | | | | | | + | |
| | | | | 15-19 (0.3) | 1.15 | | | | | | + | | + | |
| | | | | 15-19 (0.2) | 1.1 | | | + | + | | | Macular hyperpigmentation | + | |
| | | | | 15-19 (0.3) | 0.875 | | | | | | | | | |
| | | | | 15-19 (0.2) | | | | | | | | Maculopathy | + | |
| | | | | 15-19 (0.6) | 0.55 | | | | | | | | + | |
| | | | | 20-24 (1.7) | 0.95 | | | + | | + | | Maculopathy | + | |
| | | | | 20-24 (0.5) | 1 | | | + | + | + | | Optic nerve drusen | + | |
| | | | | 20-24 (1) | | Normal | | + | | + | + | | + | |
| | | | | 20-24 (0.6) | | | | | | | | | | Central foveal thickness OD 175 µm, OS 212 µm |
| | | | | 20-24 (0.6) | 1.3 | | | + | | + | | Macular pigmentary changes, Optic nerve drusen | + | |
| | | | | 20-24 (0.1) | 1.1 | | | | | | | | + | |
| | | | | 25-29 (4.5) | | | | | | | | | | Marked central peri-foveal atrophy surrounded by diffuse retinal thinning. |
| | | | | 25-29 (0.4) | | | | + | | | | | + | |
| | | | | 30-34 (2.8) | 1.3 | Normal | | | | | | | + | Interval mild enlargement of the central peri-foveal atrophy with stable adjacent diffuse retinal thinning |
| | | | | 30-34 (0.6) | | | | | | | | | + | |
| | | | | 30-34 (2.8) | | | | | | | | | + | |
| | | | *ΔT (t)* | *27.0* | | | | | | | | | | |

Supplementary Table 3.

| Patient # | Disease severity subgroup | Mutations | Sex | Age range (time since previous records) (y) | Avg VA ALL (LogMar) | Anterior segment | Fundus | | | | | | Nystagmus | OCT Description |
|---|---|---|---|---|---|---|---|---|---|---|---|---|---|---|
| | | | | | | | Normal | Abn.pigmentation in peripheral retina | Optic disc pallor | Attenuated vessels | Retinal degeneration | Fundus – Other descriptions | | |
| | | | *ΔVA* | | *0.45* | | | | | | | | | |
| | | | *ΔVA/year* | | *0.017* | | | | | | | | | |
| 9 | PEX1 p.G843D / G843D - Mild ZSD | PEX1 c.2528G>A (p.G843D) / c.2528G>A (p.G843D) | M | 0-4 | | | | | | | | | - | |
| | | | | 0-4 (0.5) | | | | + | | | | | | |
| | | | | 0-4 (1) | 0.4 | | | + | | | | | | |
| | | | | 0-4 (1.1) | 0.55 | | | + | | | | | | |
| | | | | 5-9 (1.5) | 0.85 | | | + | | | | | | |
| | | | | 5-9 (0.6) | 0.9 | | | + | + | | | | - | |
| | | | | 5-9 (0.9) | 0.675 | | | + | | | | Macular pigmentary changes | | Shows schitic like appearance of the maculae |
| | | | | 5-9 (1.4) | 0.75 | | | + | + | | | | | |
| | | | | 5-9 (0.1) | 0.65 | | | | | | | | | |
| | | | | 5-9 (0.6) | | | | | | | | | | Central foveal thickness OD 443 µm, OS 323 µm. Intra-retinal schitic spaces in both the outer nuclear layer (ONL) and inner nuclear layer (INL) that coalesce centrally. |
| | | | | 10-14 (0.5) | | | | | | | | | | Central foveal thickness OD 467 µm, OS 343 µm. Progression and the enlargement of the schitic cavities |
| | | | | 10-14 (0.9) | | | | | | | | | | Stable with significant amount of fluid. Central foveal thickness OD 459 µm, OS 324 µm. Progression of the schitic cavities. |
| | | | *ΔT (y)* | *5.6* | | | | | | | | | | |
| | | | *ΔVA* | | *0.25* | | | | | | | | | |
| | | | *ΔVA/year* | | *0.045* | | | | | | | | | |
| 10 | PEX1 p.G843D / G843D - Mild ZSD | PEX1 c.2528G>A (p.G843D) / c.2528G>A (p.G843D) | M | 0-4 | | | | | | | | | - | |
| (Patient 9's sibling) | | | | 5-9 (0.5) | 0.8 | | | + | | | | | | |
| | | | | 5-9 (1) | 1 | | | + | | | | Macular atrophy | | |
| | | | | 5-9 (0.6) | 1 | | | + | | | | Macular atrophy | - | |
| | | | | 5-9 (0.5) | 1 | | | + | | | | Macular atrophy | | |
| | | | | 5-9 (0.9) | 1 | | | | | | | | | |
| | | | | 5-9 (0.1) | 1 | | | + | | | | | | |
| | | | | 5-9 (0.5) | 1 | | | + | | + | | Macular pigmentary changes | | |
| | | | | 10-14 (0.5) | 0.875 | | | + | | | | Macular pigmentary changes | | Shows schitic like appearance of the maculae. |
| | | | | 10-14 (1.4) | | | | + | + | | | | | Central schisis in both eyes at the maculae |
| | | | | 10-14 (0.3) | 0.8 | | | + | | | | Macular atrophy | | |
| | | | | 10-14 (0.2) | | | | | | | | Central schisis in both maculae. | | Central foveal thickness OD 592 µm, OS 424 µm. Intra-retinal schitic spaces in both the outer nuclear layer (ONL) and inner nuclear layer (INL) that coalesce centrally. |
| | | | | 10-14 (0.2) | 0.75 | | | | + | | | | | |

Supplementary Table 3.

| | | | | | | | Fundus | | | | | | | |
|---|---|---|---|---|---|---|---|---|---|---|---|---|---|---|
| **Patient #** | **Disease severity subgroup** | **Mutations** | **Sex** | **Age range (time since previous records) (y)** | **Avg VA ALL (LogMar)** | **Anterior segment** | Normal | Abn.pigmentation in peripheral retina | Optic disc pallor | Attenuated vessels | Retinal degeneration | Fundus – Other descriptions | **Nystagmus** | **OCT Description** |
| | | | | 10-14 (0.7) | | | | | | | | | | Central foveal thickness OD 139 µm, OS 643 µm. Progression of the schitic cavities |
| | | | | 10-14 (0.7) | 0.75 | | | | | | | | | Significant fluid and disruption of both maculae. Progression of the schitic cavities. Central foveal thickness OD 435 µm, OS 620 µm |
| | | *ΔT (t)* | | *8.6* | | | | | | | | | | |
| | | *ΔVA* | | | *-0.05* | | | | | | | | | |
| | | *ΔVA/year* | | | *-0.006* | | | | | | | | | |
| 11 | PEX1 p.G843D / G843D - Mild ZSD | PEX1 c.2528G>A (p.G843D) / c.2528G>A (p.G843D) | M | 0.4 0-4 | | | | + | | | | | + | |
| | | | | 0-4 (0.2) | | | | + | | | + | Optic nerve hypoplasia, Macular pigmentary changes | + | |
| | | | | 0-4 (0.6) | | | | + | | | | RPE stripping in periphery, no distinguishable optic discs | + | |
| | | | | | | | | | | | | | + | |
| | | | | 0-4 (2.2) | | | | | | | + | | + | |
| | | | | 0-4 (0.2) | 0.45 | | | | | | | | + | |
| | | | | 5-9 (4.7) | 0.25 | | | + | | | + | | | |
| | | | | 10-14 (1.8) | | | | + | + | | | | | |
| | | | | 10-14 (0.3) | | | | + | | | + | Optic nerve hypoplasia | | |
| | | | | 10-14 (0.9) | | | | | | | | Optic nerve hypoplasia | | |
| | | | | 10-14 (0.8) | | | | | | | | | + | |
| | | | | 10-14 (2.6) | 0.7 | | | | | | | | + | |
| | | | | 15-19 (0.6) | | | | + | + | + | | | | |
| | | | | 15-19 (1.6) | 0.65 | | | + | + | | | | | |
| | | | | 15-19 (1.1) | 0.8 | | | | | | | | | |
| | | | | 20-24 (4.4) | 0.7 | | | | | | | | | |
| | | | | 20-24 (0.1) | 0.75 | | | + | | | | | | |
| | | | | 25-29 (4.1) | | | | | | | | | + | |
| | | *ΔT (y)* | | *18.9* | | | | | | | | | | |
| | | *ΔVA* | | | *0.3* | | | | | | | | | |
| | | *ΔVA/year* | | | *0.016* | | | | | | | | | |
| 12 | PEX1 p.G843D / G843D - Mild ZSD | PEX1 c.2528G>A (p.G843D) / c.2528G>A (p.G843D) | M | 0-4 | | | | | | | + | Optic atrophy, Macular degeneration | + | |
| (Patient 7's sibling) | | | | 0-4 (0.1) | | Normal | | | | + | | Macular pigmentary changes, Small optic discs | + | |
| | | | | 0-4 (0.2) | | | | | | | | | + | |
| | | | | 0-4 (0.6) | | Normal | | + | | + | | Small optic discs | + | |
| | | | | 10-14 (11.6) | 1 | | | | | | | | | |
| | | | | 15-19 (2) | 1.1 | | | + | | | | | | |
| | | | | 15-19 (3) | 2 | Aphakia OD | | | | | | | | |

Supplementary Table 3.

| Patient # | Disease severity subgroup | Mutations | Sex | Age range (time since previous records) (y) | Avg VA ALL (LogMar) | Anterior segment | Fundus: Normal | Fundus: Abn.pigmentation in peripheral retina | Fundus: Optic disc pallor | Fundus: Attenuated vessels | Fundus: Retinal degeneration | Fundus – Other descriptions | Nystagmus | OCT Description |
|---|---|---|---|---|---|---|---|---|---|---|---|---|---|---|
| | | | | 15-19 (0.8) | 1 | Aphakia OD, Normal OS | | + | + | + | | Optic atrophy | | |
| | | | | 20-24 (1.2) | 1 | Aphakia OD | | | | | | | | |
| | | | | 20-24 (3.2) | | Aphakia OD, Glaucoma | | | | | | | | |
| | | | *ΔT (y)* | *7.0* | | | | | | | | | | |
| | | | *ΔVA* | | *0* | | | | | | | | | |
| | | | *ΔVA/year* | | *0.000* | | | | | | | | | |
| 13 | PEX1 p.G843D / G843D - Mild ZSD | PEX1 c.2528G>A (p.G843D) / c.2528G>A (p.G843D) | F | 0-4 | | | | | | | | Optic nerve hypoplasia | + | |
| | | | | 0-4 (0.5) | | | | | | | | Optic nerve hypoplasia | + | |
| | | | | 0-4 (0.5) | | | | | | | | Optic nerve hypoplasia | + | |
| | | | | 0-4 (0.2) | | | | | | | | Optic nerve hypoplasia | | |
| | | | | 0-4 (0.4) | | | | | | | | | + | |
| | | | | 0-4 (0.2) | | | | | | | | Optic nerve hypoplasia | | |
| | | | | 0-4 (1.5) | | | | | | | | | + | |
| | | | | 0-4 (0.3) | | Normal | | + | | + | + | | + | |
| | | | | 5-9 (0.7) | 0.75 | | | | | | | | | |
| 14 | PEX1 p.G843D / G843D - Mild ZSD | PEX1 c.2528G>A (p.G843D) / c.2528G>A (p.G843D) | F | 0-4 | | | | + | | | + | Maculopathy | + | |
| | | | | 0-4 (0.6) | | | | | | | | | + | |
| | | | | 0-4 (0.8) | | | | | | | | | | Extensive and severe CME and severe disc edema. Intra-retinal schitic spaces in both the outer nuclear layer (ONL) and inner nuclear layer (INL) that coalesce centrally. Elevated optic nerve. |
| | | | | 0-4 (0.4) | | | | | | | | | | Extensive and severe CME and severe disc edema |
| | | | | 0-4 (0.2) | | | | | | | | | | Presence of macular schisis OU- persistent and significant. Optic nerve head height is 0.94 mm in the right at 0.7 mm in the left - similar to last visit. Extensive and severe CME and severe disc edema. Can see remnants of the ellipsoid zone (EZ) in some images |
| 15 | PEX1 p.G843D / G843D - Mild ZSD | PEX1 c.2528G>A (p.G843D) / c.2528G>A (p.G843D) | M | 15-19 | | Normal | | + | + | + | | | | |
| 16 | PEX1 p.G843D/Null - Intermediate ZSD | PEX1 c.2528G>A (p.G843D) / c.2097insT (p.I700Yfs42X) | M | 0-4 | | Atypical | | + | | + | | Small optic discs | + | |
| | | | | 0-4 (0.1) | | White webbing across pupils | | + | | | | | + | |
| | | | | 0-4 (0.4) | | | | | | | | | + | |
| | | | | 0-4 (0.2) | | | | | | | | | + | |
| | | | | 0-4 (1.1) | | | | | | | | Optic nerve atrophy | | |
| 17 | PEX1 p.G843D/Null - Intermediate ZSD | PEX1 c.2528G>A (p.G843D) / c.2926+1 G>A | F | 0-4 | | Normal | | + | | + | + | Optic nerve atrophy | - | |

Supplementary Table 3.

| Patient # | Disease severity subgroup | Mutations | Sex | Age range (time since previous records) (y) | Avg VA ALL (LogMar) | Anterior segment | Fundus | | | | | | Nystagmus | OCT Description |
|---|---|---|---|---|---|---|---|---|---|---|---|---|---|---|
| | | | | | | | Normal | Abn.pigmentation in peripheral retina | Optic disc pallor | Attenuated vessels | Retinal degeneration | Fundus – Other descriptions | | |
| | | | | 0-4 (0.6) | | | | | | | + | | - | |
| | | | | 0-4 (0.5) | | | | | | + | + | RPE detachment | - | |
| | | | | 0-4 (0.5) | | | | | | | + | | - | |
| | | | | 0-4 (0.5) | | | | | | + | + | | - | |
| | | | | 0-4 (0.6) | | Normal | | | | | | | | |
| | | | | 5-9 (1.2) | | Normal | | + | + | + | | Optic nerve atrophy | | |
| 18 | PEX1 p.G843D/Null - Intermediate ZSD | PEX1 c.2528G>A (p.G843D) / c.2097insT (p.I700Yfs42X) | F | 0-4 | | | | + | + | | | | + | |
| | | | | 0-4 (0.1) | | | | | | | | | + | |
| | | | | 0-4 (0.1) | | Stellate irises | | | | | | | + | |
| | | | | 0-4 (0.5) | | | | | | | | | + | |
| | | | | 0-4 (0.8) | | Lenses faint posterior subcapsular changes. | | + | | | | Retinal detachment | | |
| | | | | 5-9 (6.5) | | Normal | | + | + | | + | Thickening and fibrotic scarring around optic nerve | + | |
| 20 | PEX1 p.G843D/Null - Intermediate ZSD | PEX1 c.2528G>A (p.G843D) / c.2097insT (p.I700Yfs42X) | F | 0-4 | | Normal | | | | | | | | |
| | | | | 0-4 (0.7) | | | | | | | | | + | |
| 21 | PEX1 p.G843D/Null - Intermediate ZSD | PEX1 c.2528G>A (p.G843D) / c.2916delAfs19* | M | 0-4 | | | | | + | | | | + | |
| | | | | 0-4 (0.1) | | | | | | | | | + | |
| 22 | PEX1 p.G843D/Null - Intermediate ZSD | PEX1 c.2528G>A (p.G843D) / c.1109insA (p.I370NfsX2) | F | 0-4 | | | | | | | | | + | |
| | | | | 0-4 (0.2) | | | | | | | | | + | |
| | | | | 0-4 (0.5) | 0.7 | | | | | | | | + | |
| | | | | 0-4 (0.1) | | | | | | | | | + | |
| | | | | 0-4 (1.1) | | | | | | | | | - | |
| | | | | 0-4 (1.1) | 1.6 | | | | | | | Macular pigmentary changes | - | |
| | | | | 5-9 (1.1) | | | | | | | | Macular pigmentary changes, Non-tortuous retinal vessels | - | |
| | | | | 5-9 (1) | | | | | | | | | + | |
| | | *ΔT (t)* | | *2.3* | | | | | | | | | | |
| | | *ΔVA* | | | *0.9* | | | | | | | | | |
| | | *ΔVA/year* | | | *0.398* | | | | | | | | | |
| 23 | PEX1 p.G843D/Null - Intermediate ZSD | PEX1 c.2528G>A (p.G843D) / p.Asn1088fs | M | 15-19 | | | | | | | | | + | |
| 24 | PEX1 p.G843D/Null - Intermediate ZSD | PEX1 c.2528G>A (p.G843D) / c.1108delA | M | 0-4 | | | | | + | | | Optic nerve atrophy | + | |
| | | | | 0-4 (0.5) | | | | | | | | | + | |
| | | | | 0-4 (0.2) | | | | | | | | | + | |

Supplementary Table 3.

| Patient # | Disease severity subgroup | Mutations | Sex | Age range (time since previous records) (y) | Avg VA ALL (LogMar) | Anterior segment | Fundus: Normal | Fundus: Abn.pigmentation in peripheral retina | Fundus: Optic disc pallor | Fundus: Attenuated vessels | Fundus: Retinal degeneration | Fundus – Other descriptions | Nystagmus | OCT Description |
|---|---|---|---|---|---|---|---|---|---|---|---|---|---|---|
| | | | | 0-4 (0.6) | | Normal | | | | | | | + | |
| | | | | 0-4 (0.2) | | Normal | | | | | | Optic nerve atrophy | + | |
| | | | | 0-4 (0.3) | | Normal | | | | | | Optic nerve atrophy | + | |
| 25 | PEX1 p.G843D/Null - Intermediate ZSD | PEX1 c.2528G>A (p.G843D) / c.382C>T (p.Q128X) | M | 0-4 | | Normal | | | | | | | | |
| | | | | 0-4 (0.3) | | Normal | | | | | | | | |
| | | | | 0-4 (0.2) | 0.9 | Normal | | | | | | Hypopigmented spot in the superotemporal macula OD | | |
| | | | | 0-4 (1.1) | | | | | | | | | + | |
| | | | | 0-4 (1.2) | | | | | | | | | - | |
| | | | | 0-4 (1.5) | | Normal | | + | | | | Optic nerve OD is obstructed with a fibrous stalk, Macular pigmentary changes | | |
| | | | | 5-9 (0.5) | | | | + | + | + | + | Optic nerve atrophy | | |
| | | | | 5-9 (0.5) | | Normal | | | | | | Gliotic material on the optic discs OU, Macular hyperpigmentation | | |
| | | | | 5-9 (1.2) | | Normal | | | | | | Gliotic material on optic discs | | |
| 26 | PEX1 p.G843D/Null - Intermediate ZSD | PEX1 c.2528G>A (p.G843D) /c.2097insT (p.I700Yfs42X) | M | 10-14 | | | | + | | | | Optic nerve hypoplasia | | |
| | | | | 15-19 (2.7) | | | | | | | | | - | |
| | | | | 15-19 (0.2) | | | | | | | | Optic nerve atrophy | - | |
| 27 | PEX1 p.G843D/Null - Intermediate ZSD | PEX1 c.2528G>A (p.G843D) /c.2097insT (p.I700Yfs42X) | M | 5-9 | | | | + | + | + | + | | + | |
| | | | | 15-19 (8.7) | | Normal | | | | | + | | + | |
| | | | | 15-19 (0.1) | | Normal | | + | + | + | | Macular pigmentary changes, Small optic disc | | |
| | | | | 20-24 (2.9) | | Cataracts | | | | | | | + | |
| | | | | 20-24 (1.5) | | Cataracts | | | | | | | + | |
| 28 | PEX1 p.G843D/Null - Intermediate ZSD | PEX1 c.2528G>A (p.G843D) /c.2097insT (p.I700Yfs42X) | F | 0-4 | | | | | | | | | + | |
| | | | | 0-4 (0.2) | | | | | | | | | + | |
| | | | | 0-4 (0.2) | | | | + | | | | | + | |
| | | | | 0-4 (0.2) | | | | | | | | | + | |
| | | | | 0-4 (1) | | | | | | | | Retina scarring | + | |
| | | | | 0-4 (1) | | | | | | | | | + | |
| 29 | PEX1 p.G843D/Null - Intermediate ZSD | PEX1 c.2528G>A (p.G843D) /c.2097insT (p.I700Yfs42X) | F | 0-4 | | | | + | | | | | + | |
| 30 | PEX1 p.G843D/Null - Intermediate ZSD | PEX1 c.2528G>A (p.G843D) /c.2097insT (p.I700Yfs42X) | M | 10-14 | | | | | + | | | | | |
| 31 | PEX1 p.G843D/Null - Intermediate ZSD | PEX1 c.2528G>A (p.G843D) / c.2916delA (p.G973AfsX16) | M | 0-4 | | | | | | | + | | | |

Supplementary Table 3.

| Patient # | Disease severity subgroup | Mutations | Sex | Age range (time since previous records) (y) | Avg VA ALL (LogMar) | Anterior segment | Fundus: Normal | Fundus: Abn.pigmentation in peripheral retina | Fundus: Optic disc pallor | Fundus: Attenuated vessels | Fundus: Retinal degeneration | Fundus – Other descriptions | Nystagmus | OCT Description |
|---|---|---|---|---|---|---|---|---|---|---|---|---|---|---|
| 32 | PEX1 p.G843D/Null - Intermediate ZSD | PEX1 c.2528G>A (p.G843D) /c.2097insT (p.I700Yfs42X) | F | 0-4 | | | | | | | | | + | |
| | | | | 0-4 (1.2) | | | | | | | | | + | |
| 33 | PEX1 p.G843D/Null - Intermediate ZSD | PEX1 c.2528G>A (p.G843D) /c.2097insT (p.I700Yfs42X) | F | 0.5 0-4 | | | | + | | | | Optic disc swelling | + | |
| | | | | 0-4 (0.5) | | | | | | | | | + | |
| | | | | 0-4 (2.5) | 1.6 | | | + | | | | | | |
| | | | | 0-4 (1.1) | 1.6 | | | | | | | | | |
| | | | | 5-9 (1.2) | 1.3 | | | | | | | | | |
| | | | | 5-9 (0.6) | 1.2 | | | | | | | | | |
| | | | | 5-9 (0.8) | | | | | | | | | + | |
| | | | | 5-9 (0.6) | | Normal | | + | | | | | + | |
| | | | | 5-9 (0.7) | | Normal | + | + | | | | | + | |
| | | | | 10-14 (1) | | | | + | | + | | Optic atrophy | + | |
| | | | | 10-14 (2) | 1 | | | + | | | | | | |
| | | | | 10-14 (0.5) | | | + | | | | | | | |
| | | | | 10-14 (0.7) | | | | + | + | | | | | |
| | | | | 10-14 (0.9) | | | | | + | | | | + | |
| | | | | 15-19 (0.8) | | | | + | | | | | + | |
| | | | | 15-19 (0.9) | | | | + | + | | | | | |
| | | | | 15-19 (1.9) | | | | | | | | | + | |
| | | | | 15-19 (0.6) | | | | + | | | | | | |
| | | | | 20-24 (1.4) | | Left vitreous asteroid hyalosis. Small cataract OD | | + | + | + | | | - | |
| | | | | 20-24 (3) | | Asteroid hyalosis | | | | | | Optic atrophy | | |
| | | | | 35-39 (12) | | | | | | | | | + | |
| | | | *ΔT (t)* | *9.0* | | | | | | | | | | |
| | | | *ΔVA* | | *-0.6* | | | | | | | | | |
| | | | *ΔVA/year* | | *-0.067* | | | | | | | | | |
| 34 | PEX1 p.G843D/Null - Intermediate ZSD | PEX1 c.2528G>A (p.G843D) /c.2097insT (p.I700Yfs42X) | F | 0-4 | | | | | | | | | - | |
| | | | | 0-4 (0.2) | | | + | | | | | | | |
| | | | | 0-4 (0.1) | | | | + | | | | | | |
| | | | | 0-4 (0.5) | | | | | | | | | + | |
| | | | | 15-19 (18) | | Inferocentral cornea dry w mild epithelia erosion inferiorly OU | | + | + | + | + | | | |
| 35 | PEX1 p.G843D/Null - Intermediate ZSD | PEX1 c.2528G>A (p.G843D) / c.2383C>T (p.Arg795fs*) | M | 0-4 | | | | | | | | | + | |
| | | | | 0-4 (2.2) | | | | | | | | | + | |

Supplementary Table 3.

| Patient # | Disease severity subgroup | Mutations | Sex | Age range (time since previous records) (y) | Avg VA ALL (LogMar) | Anterior segment | Fundus | | | | | | Nystagmus | OCT Description |
|---|---|---|---|---|---|---|---|---|---|---|---|---|---|---|
| | | | | | | | Normal | Abn.pigmentation in peripheral retina | Optic disc pallor | Attenuated vessels | Retinal degeneration | Fundus – Other descriptions | | |
| | | | | 15-19 (13.8) | | Normal | | + | + | + | | Optic atrophy | | |
| | | | | 15-19 (2.1) | | Normal | | + | + | | | Optic atrophy | - | |
| 19 | Mild-Intermediate ZSD | PEX1 c.2528G>A (p.G843D) / c.1961insCAGTGTGGA (p.Trp653_met654insThrVal Trp) | F | 0-4 | | | | | | | | Small optic nerve | - | |
| | | | | 5-9 (5) | | Normal | | | | | | | | |
| 36 | Mild-Intermediate ZSD | PEX1 c.1642C>T (p.L548F) / c.1642C>T (p.L548F) | M | 0-4 | | | | + | | + | | | | |
| | | | | 5-9 (1.7) | | Normal | | | | | + | | + | |
| | | | | 5-9 (0.2) | | | + | | | + | | | + | |
| | | | | 5-9 (0.2) | | | | | | | | | + | |
| | | | | 5-9 (1.1) | | Normal | | | | + | | Macular atrophy | + | |
| | | | | 5-9 (1.9) | | | | | | + | | | + | |
| 37 | Mild-Intermediate ZSD | PEX6 c.659G>T (p.G220V) / c.2095-22_2095-11delCCACGCACTTTC | M | 0-4 | | ` | | | | | + | | + | |
| | | | | 0-4 (1.6) | | | | | | | | | + | |
| | | | | 0-4 (0.1) | | | | | + | | + | Optic nerve atrophy | + | |
| | | | | 0-4 (0.1) | | | | + | | | | Macular pigmentary changes | | |
| | | | | 0-4 (0.2) | | | + | | | | | | + | |
| | | | | 0-4 (0.4) | | | | | | | + | | + | |
| | | | | 0-4 (0.4) | 0.85 | | | + | | | + | Indistinct fovea | + | |
| | | | | 5-9 (1) | 0.925 | Normal | | | | | | | + | |
| | | | | 5-9 (1) | 0.85 | Normal | | | | | + | Indistinct fovea | + | |
| | | | | 5-9 (1.1) | 0.85 | Normal | | + | + | + | + | | + | |
| | | | | 5-9 (0.9) | 0.85 | | | | | | + | | | |
| | | | *ΔT (t)* | *4.0* | | | | | | | | | | |
| | | | *ΔVA* | | *0* | | | | | | | | | |
| | | | *ΔVA/year* | | *0.000* | | | | | | | | | |
| 38 | Mild-Intermediate ZSD | PEX1 c.2916delA (p.G973fs) / c.1777G>A (p.G593R) | M | 0-4 | | Normal | | | | | + | Flat macula | + | |
| | | | | 0-4 (0.2) | | | | | | | | | + | |
| | | | | 0-4 (0.6) | | | | | | | | | + | |
| 39 | Mild-Intermediate ZSD | PEX6 c.2578C>T (p.R860W) / + | M | 0-4 | | Normal | + | | | | | | - | |
| | | | | 0-4 (0.5) | | | + | | | | | | - | |
| 40 | Mild-Intermediate ZSD | PEX1 c.1642C>T (p.L548F) / c.1642C>T (p.L548F) | F | 0-4 | | Normal | + | | | | | | | |
| (Patient 36's sibling) | | | | 0-4 (0.7) | | | | | | | | | + | |

Supplementary Table 3.

| Patient # | Disease severity subgroup | Mutations | Sex | Age range (time since previous records) (y) | Avg VA ALL (LogMar) | Anterior segment | Fundus | | | | | | Nystagmus | OCT Description |
|---|---|---|---|---|---|---|---|---|---|---|---|---|---|---|
| | | | | | | | Normal | Abn.pigmentation in peripheral retina | Optic disc pallor | Attenuated vessels | Retinal degeneration | Fundus – Other descriptions | | |
| | | | | 0-4 (1.4) | | | | | | | | | - | |
| 41 | Mild-Intermediate ZSD | PEX5 c.1803_1805delGGAinsTC (p.E601Dfs14*) / c. 1799C>T (p.S600L) | M | 5-9 | | | | | | | | | - | |
| | | | | 5-9 (0.1) | | Normal | + | | | | | | - | |
| | | | | 5-9 (0.2) | | | | | | | | | - | |
| | | | | 5-9 (0.3) | | | | | | | | | + | |
| 42 | Mild-Intermediate ZSD | PEX1 c.2097dupT (p. I700fs*) / del exon24 and 3'UTR | M | 0-4 | | | | | | | | | + | |
| | | | | 0-4 (2) | | | | | | | + | | | |
| 43 | Mild-Intermediate ZSD | PEX6 c.2125G>A (p.Gly709Arg) / c.2125G>A (p.Gly709Arg) | M | 0-4 | | Normal | + | | | | | | - | |
| | | | | 0-4 (0.3) | | Normal | | | | | | | - | |
| | | | | 0-4 (0.4) | | Normal | | | | | | | | |
| | | | | 0-4 (0.7) | | Normal | + | | | | | | - | |
| | | | | 0-4 (0.2) | | Normal | | | | | | | | |
| | | | | 0-4 (0.4) | 0.65 | | | | | | | | | |
| | | | | 0-4 (0.6) | | Normal | | | | | | | - | |
| 44 | Mild-Intermediate ZSD | PEX6 c.2125G>A (p.Gly709Arg) / c.2125G>A (p.Gly709Arg) | F | 0-4 | | | + | | | | | | - | |
| (Patient 43's sibling) | | | | 0-4 (0.3) | | Normal | | | | | | | - | |
| | | | | 0-4 (0.6) | | Normal | | | | | | | | |
| | | | | 0-4 (1.6) | | Normal | + | | | | | | | |
| 45 | Mild-Intermediate ZSD | PEX26 c.292C>T (p.R98W) / c.292C>T (p.R98W) | M | 0-4 | | | | + | | | | | + | |
| | | | | 0-4 (0.3) | | | | | | | | | - | |
| | | | | 0-4 (0.3) | | | | + | | | | | | |
| | | | | 10-14 (6.9) | 1.05 | Unusual lacy and granular corneal opacity in anterior stroma OU | | + | | | | | | |
| | | | | 10-14 (0.4) | | Band keratinopathy OU | | + | | | | | | |
| | | | | 10-14 (1.2) | | Bilateral uveitis with band keratinopathy OU | | + | | | | Macular pigmentary changes | | |
| 46 | Mild-Intermediate ZSD | PEX12 c.1–26G>A; c.102A>T (p.Arg34Ser) / c.1–26G>A; c.102A>T (p.Arg34Ser) | M | 0-4 | | | | | + | | | | | |

Supplementary Table 3.

| Patient # | Disease severity subgroup | Mutations | Sex | Age range (time since previous records) (y) | Avg VA ALL (LogMar) | Anterior segment | Fundus: Normal | Fundus: Abn.pigmentation in peripheral retina | Fundus: Optic disc pallor | Fundus: Attenuated vessels | Fundus: Retinal degeneration | Fundus – Other descriptions | Nystagmus | OCT Description |
|---|---|---|---|---|---|---|---|---|---|---|---|---|---|---|
| | | | | 0-4 (4.2) | | Exposure keratopathy OU. Corneal abrasion OS. | + | | | | | | | |
| | | | | 5-9 (0.6) | | Exposure keratopathy OU. Corneal abrasion OS. | + | | | | | Optic atrophy | - | |
| 47 | Mild-Intermediate ZSD | PEX1 c.2916delA (p.G973AfsX16) / c.484C>A (p.P162T) | M | 0-4 | | | | + | | | | | + | |
| 48 | Mild-Intermediate ZSD | PEX1 c.547G>A (p.Arg183Ter) / c.1099delG (p.Gln367fs*) | M | 0-4 | | | | | | | | | + | |
| 49 | Mild-Intermediate ZSD | PEX1 c.547G>A (p.Arg183Ter) / c.1099delG (p.Gln367fs*) | M | 5-9 | | | | | | | | | + | |
| (Patient 48's sibling) | | | | 5-9 (0.1) | | Central corneal opacities | | | | | + | Abnormal optic nerve | + | |
| 50 | Mild-Intermediate ZSD | PEX26 c.35dupC (p.L12Pfs) / c.292C>T (p.Arg98Trp) | M | 0-4 | | Whitish deposits in posterior side of vitreous humour OU | | + | | | | Flat detachment of the retina | | |
| | | | | 0-4 (1.1) | | | | | | | | | - | |
| 51 | Mild-Intermediate ZSD | PEX1 c.2528G>A (p.G843D) / c.2528G>A (p.G843D) ; PEX6 c.1802G>A (p.Arg601Gln) / c.1637G>A (p.Arg546His) | M | 0-4 | | | | + | | | | | + | |
| | | | | 0-4 (0.4) | | | | | | | + | | | |
| 52 | Mild-Intermediate ZSD | PEX6 c.10-69del, 126-217del (p.Gly44Thrfs*3) / c.331delG | F | 0-4 | | Normal | + | | | | | | | |
| | | | | 0-4 (0.1) | | Normal | | | + | | | Small optic nerve | | |
| | | | | 0-4 (0.1) | | Normal | + | | | | | | | |
| | | | | 0-4 (0.2) | | | | + | | | + | | + | |
| | | | | 0-4 (0.2) | | | | | | | + | | | |
| | | | | 0-4 (0.8) | | | | | | | | | + | |
| | | | | 0-4 (2) | | | | | | | + | | + | |
| 53 | Mild-Intermediate ZSD | PEX6 c.2837_2839del (p.L946_T947delinsP) / c.2837_2839del (p.L946_T947delinsP) | F | 0-4 | | | | | | | | | + | |
| 54 | Mild-Intermediate ZSD | PEX1 c.2097dupT (p. I700fs*) / del exon24 and 3'UTR | F | 0-4 | | | | | | | | | + | |
| | | | | 0-4 (1.2) | | | | | | | | | + | |
| | | | | 0-4 (0.4) | | Normal | | | | | | | | |
| | | | | 0-4 (0.2) | | Normal | | | | | | | | |

Supplementary Table 3.

| Patient # | Disease severity subgroup | Mutations | Sex | Age range (time since previous records) (y) | Avg VA ALL (LogMar) | Anterior segment | Fundus | | | | | | Nystagmus | OCT Description |
|---|---|---|---|---|---|---|---|---|---|---|---|---|---|---|
| | | | | | | | Normal | Abn.pigmentation in peripheral retina | Optic disc pallor | Attenuated vessels | Retinal degeneration | Fundus – Other descriptions | | |
| | | | | 0-4 (0.3) | | Normal | | | | | | Macular pigmentary changes, no discernable optic disc | + | |
| 55 | Mild-Intermediate ZSD | PEX19 c.764T>A (M225K) / c.764T>A (M225K) | F | 0-4 | | | | + | | | | Optic nerve atrophy | | |
| 56 | Mild-Intermediate ZSD | PEX16 c.692A>G (p.His231Arg) / c.692A>G (p.His231Arg) | M | 5-9 | | Normal | + | | | | | | | |
| | | | | 10-14 (5) | 0.725 | Cataracts | + | | | | | | | |
| 57 | Mild-Intermediate ZSD | PEX6 c.1220C>A (p.T407N) / c.1220C>A (p.T407N) | M | 0-4 | | Normal | + | | | | | | - | |
| | | | | 0-4 (0.2) | | Normal | | | | | | | | |
| | | | | 0-4 (0.8) | | Normal | | | + | | | Optic nerve atrophy | | |
| | | | | 0-4 (0.3) | | Normal | | | | | | Optic nerve atrophy | - | |
| 58 | Mild-Intermediate ZSD | PEX16 c.372delG (p.Arg124fs) / c.683C>T (p.Pro228Leu) | M | 5-9 | | Normal | + | | | | | | | |
| | | | | 10-14 (2) | | Normal | + | | | | | | | |
| 59 | Mild-Intermediate ZSD | PEX16 c.372delG (p.Arg124fs) / c.683C>T (p.Pro228Leu) | F | 0-4 | | Normal | + | | | | | | | |
| (Patient 58's sibling) | | | | 0-4 (0.2) | | Normal | + | | | | | | | |
| | | | | 0-4 (1) | | Normal | + | | | | | | | |
| | | | | 5-9 (0.7) | | Normal | | | + | | | | | |
| 60 | Mild-Intermediate ZSD | PEX1 c.2528G>A (p.G843D) / c.439G>T (p.V147F) | M | 0-4 | | | | + | | | | | | |
| | | | | 0-4 (0.1) | | | | | | | | | + | |
| | | | | 5-9 (3.5) | 0.875 | | | + | | + | | | | |
| | | | | 5-9 (0.2) | | | | + | + | + | | | | |
| | | | | 5-9 (1.4) | 0.8 | | | + | | | | | | CME. Starting CAI eye drops (Brinzolamide). |
| | | | | 5-9 (0.1) | 0.65 | | | | | | | | | CME |
| | | | | 5-9 (0.1) | 0.75 | | | + | | | | | | CME |
| | | | | 5-9 (0.6) | 0.7 | | | | | | | | | CME. Stopped CAI eye drops (Brinzolamide) |
| | | | | 5-9 (0.9) | 0.7 | | | | | | | | | CME, less macular edema. Central foveal thickness OD 288 µm, OS 273 µm |
| | | | *ΔT (y)* | *3.3* | | | | | | | | | | |
| | | | *ΔVA* | | *-0.175* | | | | | | | | | |
| | | | *ΔVA/year* | | *-0.053* | | | | | | | | | |
| 61 | Mild-Intermediate ZSD | PEX1 c.2966T>C (p.Ile989Thr) / PEX1 c.2097dupT (p.I700Yfs42X) | M | 0-4 | | Normal | | + | | | | | + | |
| | | | | 0-4 (0.6) | | | | | | | | | + | |
| | | | | 0-4 (0.4) | | | | | | | | | | OS: Small CME but “almost negligible”. Lot of photoreceptors still present. |

Supplementary Table 3.

| Patient # | Disease severity subgroup | Mutations | Sex | Age range (time since previous records) (y) | Avg VA ALL (LogMar) | Anterior segment | Fundus | | | | | | Nystagmus | OCT Description |
|---|---|---|---|---|---|---|---|---|---|---|---|---|---|---|
| | | | | | | | Normal | Abn.pigmentation in peripheral retina | Optic disc pallor | Attenuated vessels | Retinal degeneration | Fundus – Other descriptions | | |
| 62 | Severe ZSD | PEX26 c.173G>A (Cys58Tyr) / c.173G>A (Cys58Tyr) | M | 0-4 | | Microspherophakia | | | | | | | - | |
| 63 | Severe ZSD | PEX6 c.2398_2417del20insT / c.2094G>T | M | 0-4 | | | | + | | | | | | |
| 64 | Severe ZSD | PEX12 c.533_535delAAC (p.Q178del) / c.888_889delCT (p.L297Tfs*12) | F | 0-4 | | Normal | | | + | | | Blond periphery, small optic discs, flat and dry macula | - | |
| | | | | 0-4 (0.3) | | Normal | | | | | | Small optic discs | | |
| 65 | Severe ZSD | PEX1 c.2097insT (p.I700Yfs42X) / c.2719G>T (p.Gly907Trp) | F | 0-4 | | Mild cataract | | | | | | | + | |
| | | | | 0-4 (0.3) | | | | | | | | | + | |
| 66 | Severe ZSD | PEX10 c.874-875delCT (p.Leu292fs) / c.874-875delCT (p.Leu292fs) | F | 0-4 | | Glaucoma | + | | | | | | | |
| | | | | 0-4 (0.3) | | Cataracts | | + | | + | | | - | |
