## Supplementary Table 4 for "Peroxisome Biogenesis Disorders in the Zellweger Spectrum: Ophthalmic Findings from a New Natural History Study Cohort and Scoping Literature Review"

**Supplementary Table 4. Ophthalmic findings in previously reported Zellweger Spectrum Disorder patients.** Empty cells represent non available data. Presence of the finding is represented by "+" and absence of the finding is represented by "-". VA is OU or average of OS and OD if OU not available.
Abbreviations: CMT = Central Macular Thickness; ERG = Eletroretinogram; ERM = Epiretinal membrane; F+F = Fix and Follow; ILM = Inner nuclear layer; LP = Light perception; OCT = Optical Coherence Tomography; ONL = Outer nuclear layer; OPL – Outer Plexiform Layer; PR = Photoreceptors; RPE = Retinal pigment epithelium, VA = Visual Acuity; VEP = Visual Evoked Potential.

| **Reported Patient #** | **Disease severity subgroup** | **Mutations** | **Sex** | **Age (yr)** | **VA (LogMar)** | **Anterior segment** | **Fundus** | | | | | **Fundus – Other descriptions** | **Nystagmus** |
|---|---|---|---|---|---|---|---|---|---|---|---|---|---|
| | | | | | | | Normal | Abnormal pigmentation in peripheral retina | Optic disc pallor | Attenuated vessels | Retinal degeneration | | |
| 1[1] | Mild-Intermediate ZSD | | | | LP | | | | | | | Macular dystrophy | - |
| 2[1] | Mild-Intermediate ZSD | | | | LP | | + | | | | | | - |
| 3[1] | Mild-Intermediate ZSD | | | | LP, F+F | | | | | | + | Retinal fold | - |
| 4[1] | Mild-Intermediate ZSD | | | | 0.50 | | | + | | | + | | - |
| 5[1] | Mild-Intermediate ZSD | | | 4.3 | 0.50 | | | + | | | + | Maculopathy | - |
| 6[1] | Mild-Intermediate ZSD | | | 6 | 0.50 | | + | | | | | | - |
| 7[1] | Mild-Intermediate ZSD | | | | LP, F+F | | | | | | + | | - |
| 8[1] | Mild-Intermediate ZSD | | | 1.5 | LP, F+F | | | + | | | | Small optic discs | - |
| 9[1] | Mild-Intermediate ZSD | | | 0.9 | LP, F+F | | | + | | | | Small optic discs | - |
| 10[1] | Mild-Intermediate ZSD | | | 9 | LP, F+F | | | | | | + | Macular dystrophy | - |
| 11[1] | Mild-Intermediate ZSD | | | 7 | 1.00 | | | + | | | + | | - |
| 12[1] | Mild-Intermediate ZSD | | | | 1.00 | | | | | | + | Macular dystrophy | - |
| 13[1] | Mild-Intermediate ZSD | | | | LP | | | | | | + | Macular dystrophy | - |
| 14[2] | PEX1 p.G843D/G843D - Mild ZSD | PEX1 p.G843D / G843D | | 35 | 0.50 | | | + | | | | | |

Supplementary Table 4.

| | | | | | | | | | | | | | |
|---|---|---|---|---|---|---|---|---|---|---|---|---|---|
| 15[2] | PEX1 p.G843D/G843D - Mild ZSD | PEX1 p.G843D / G843D | | 32 | 1.30 | | | + | | | | | + |
| 16[2] | PEX1 p.G843D/G843D - Mild ZSD | PEX1 p.G843D / G843D | | 31 | 1.10 | | | + | | | | | |
| 17[2] | Mild-Intermediate ZSD | PEX11β c.64C>T / c.64C>T | | 27.5 | 1.00 | Cataracts | | | | | | | |
| 18[2] | Mild-Intermediate ZSD | PEX26 p.R98W / R98W | | 23 | 1.00 | Cataracts | | + | | | | | |
| 19[2] | PEX1 p.G843D/G843D - Mild ZSD | PEX1 p.G843D / G843D | | 22 | 1.00 | | | + | | | | | + |
| 20[2] | Mild-Intermediate ZSD | PEX1 c.1777G>A / c.2071+1G>T | | 19.5 | 1.00 | | | + | | | | | + |
| 21[2] | PEX1 p.G843D/G843D - Mild ZSD | PEX1 p.G843D / G843D | | 19 | 0.70 | | | + | | | | | |
| 22[2] | PEX1 p.G843D/G843D - Mild ZSD | PEX1 p.G843D / G843D | | 17.5 | 0.60 | | | + | | | | | |
| 23[2] | Mild-Intermediate ZSD | PEX6 p.R601Q / E664D | | 17.5 | 0.60 | | | + | | | | | + |
| 24[2] | PEX1 p.G843D/G843D - Mild ZSD | PEX1 p.G843D / G843D | | 17 | 0.60 | | | + | | | | | |
| 25[2] | Mild-Intermediate ZSD | PEX26 p.R98W / R98W | | 16 | 0.80 | | | + | | | | | + |
| 26[2] | Mild-Intermediate ZSD | PEX1 p.G843D / ? | | 28 | 0.70 | | | | | | | | |
| 27[2] | PEX1 p.G843D/G843D - Mild ZSD | PEX1 p.G843D / G843D | | 27 | 1.30 | | | + | | | | | |
| 28[2] | PEX1 p.G843D/G843D - Mild ZSD | PEX1 p.G843D / G843D | | 24.5 | 1.40 | | | | | | | | |
| 29[2] | Mild-Intermediate ZSD | PEX1 p.G843D / ? | | 22 | 1.00 | | | + | | | | | |
| 30[2] | Mild-Intermediate ZSD | PEX1 p.G843D / L879S | | 18.5 | 1.00 | | | + | | | | | + |
| 31[2] | PEX1 p.G843D/Null - Intermediate ZSD | PEX1 p.G843D / I700Yfs42X | | 18 | 1.30 | | | + | | | | | + |

Supplementary Table 4.

| | | | | | | | | | | | | | |
|---|---|---|---|---|---|---|---|---|---|---|---|---|---|
| 32[2] | PEX1 p.G843D/G843D - Mild ZSD | PEX1 p.G843D / G843D | | 17 | | | | + | | | | | + |
| 33[3] | Severe ZSD | | M | 4 do | | Thickened cornea<br>Shallow anterior segment<br>Posterior subcapsular cataracts | | | | | + | Hypercellular optic disc<br>Thinning and PR loss in posterior pole and perimacular area. | |
| 34[4] | Severe ZSD | | F | 0.06 | LP | Normal | | + | | + | + | Red lesion 1/3 of disc diameter in macula OD<br>Atrophic flattening of the RPE<br>Almost total loss of the PR outer segments | + |
| 35[5] | Severe ZSD | | F | 1.2 | | White vitreous opacities | | + | | | + | Optic disc atrophy<br>Macular degeneration | |
| 36[6] | Severe ZSD | | F | 0.6 | F+F | Diffusely cloudy appearance of cornea OD<br>Central corneal ulcer OS<br>2-3 mm hypopyon OS | + | | | | | | |
| 37[7] | Severe ZSD | | M | 0.2 | | Zonular cataracts | | | | | | | |
| 38[7] | Severe ZSD | | F | 0.19 | | Zonular cataracts | | | | | | | |
| 39[7] | Severe ZSD | | F | 0.3 | | Zonular cataracts | | + | | | | Macular degeneration<br>Optic disc atrophy | |
| 40[7] | Severe ZSD | | F | 0.8 | | Zonular cataracts | | + | | | | Macular degeneration<br>Optic disc atrophy | |
| 41[8] | Severe ZSD | | | | | | | + | | | | | + |
| 42[8] | Severe ZSD | | | | | | | + | | | | | + |
| 43[8] | Mild-Intermediate ZSD | | | | | | | + | | | | | + |
| 44[8] | Mild-Intermediate ZSD | | | | | Cataracts | | | | | | | + |
| 45[9] | Mild-Intermediate ZSD | | M | 1.5 | | Normal | | + | + | + | | | |
| 46[10] | Severe ZSD | | | 0.5 | | | | + | + | + | | | |
| 47[10] | Severe ZSD | | | 1.5 | | | | + | + | | | | |
| 48[11] | Mild-Intermediate ZSD | | M | 0.25 | | | | + | | | | | |
| 49[11] | Mild-Intermediate ZSD | | M | 4 | | Cloudy cornea | | + | | + | | | + |

Supplementary Table 4.

| | | | | | | | | | | | | | |
|---|---|---|---|---|---|---|---|---|---|---|---|---|---|
| 50[11] | Mild-Intermediate ZSD | | M | 0.4 | | | | + | | | | | + |
| 51[12] | Mild-Intermediate ZSD | PEX1 p.G843D/? | F | 5.5 | | | | + | | | + | Optic disc papilledema OU | + |
| 52[13] | PEX1 p.G843D/Null - Intermediate ZSD | PEX1 p.G843D / I700Yfs42X | F | 2 | F+F | | | + | | | | Normal optic disc<br>Large chorioretinal folds with fibrosis OD<br>ERM overlying fovea OS | + |
| 53[14] | Mild-Intermediate ZSD | | M | 1.7 | F+F | Clear corneas, lamellar/sutural cataracts OU | | + | | | + | Macula dull without foveal light reflex, chorioretinal atrophy | |
| 54[15] | Mild-Intermediate ZSD | | M | 3.6 | | | | + | + | | | | + |
| 55[15] | Mild-Intermediate ZSD | | F | 3.7 | F+F | Normal | | + | | | | Diffusely hypopigmented macula<br>Wrinkling of the ILM<br>Normal optic discs | - |
| 56[16] | Mild-Intermediate ZSD | | M | 0.6 | | Normal | | + | | + | | Hypopigmented macula<br>Grey optic discs with edematous papillae | |
| 57[17] | Mild-Intermediate ZSD | | M | 12 | | | | + | | | + | Atrophy of all retinal layers<br>Loss of rods and cones | |
| 58[18] | Mild-Intermediate ZSD | | M | 12 | 1.00 | | | + | + | + | | | - |
| 59[18] | Mild-Intermediate ZSD | | M | 15 | 1.00 | | | + | + | + | | | - |
| 60[19] | Mild-Intermediate ZSD | PEX1 compound heterozygote | M | 0.4 | | | | + | | + | | Hypoplastic choroid | + |
| 61[20] | PEX1 p.G843D/G843D - Mild ZSD | PEX1 p.G843D / G843D | F | 28 | 1.30 | | | + | + | + | | Maculopathy<br>RPE atrophy<br>Optic disc drusen | + |
| 62[21] | PEX1 p.G843D/G843D - Mild ZSD | PEX1 p.G843D / G843D | F | 25 | 0.65 | Early subcapsular cataracts OS | | + | | + | + | Optic disc drusen | |
| 63[22] | PEX1 p.G843D/p.G843D - Mild ZSD | PEX1 p.G843D / G843D | M | 8 | 0.69 | | | + | | | | Optic nerve hypoplasia | |
| 64[23] | Mild-Intermediate ZSD | PEX26 p.D43H / R98W | M | 1.8 | | | | + | | | | Macular involvment | |
| 65[24] | Severe ZSD | PEX6 p.T572I / IVS10+2T>C | M | 1 | | | | + | | | | Small optic discs | |

Supplementary Table 4.

| | | | | | | | | | | | | | |
|---|---|---|---|---|---|---|---|---|---|---|---|---|---|
| 66[24] | Mild-Intermediate ZSD | PEX6 p.T572I / T572I | M | | | | | | | | | | |
| 67[24] | Mild-Intermediate ZSD | PEX6 IVS10+2T>C / p.A809V + I845T | F | | | | | | | | | | |
| 68[25] | Mild-Intermediate ZSD | | F | 29 | 0.65 | | | + | | | | | |
| 69[26] | Mild-Intermediate ZSD | PEX1 p.L1047P / L1047P | F | 13 | | Normal | | + | + | + | | Decreased macular reflection with edema | + |
| 70[27] | Mild-Intermediate ZSD | PEX6 p.R601Q / R99L | F | 7 | | | | | | + | | Retinal dystrophy Maculopathy | |
| 71[27] | Mild-Intermediate ZSD | PEX6 p.R601Q / V92G | M | 12 | | | | | | | | Retinal dystrophy | |
| 72[28] | Mild-Intermediate ZSD | PEX6 p.R601Q / V92G | M | 12 | 1.55 | Lens cortical flecks OU | | + | | | | Normal optic disc Normal retinal vessels | |
| 73[28] | Mild-Intermediate ZSD | PEX6 p.R601Q / R99L | F | 7 | 0.00 | Lens cortical flecks OU | | + | | | | Maculopathy Normal optic disc Normal retinal vessels | |
| 74[29] | Mild-Intermediate ZSD | PEX1 p.S555P / p.I700Yfs42X | F | 30 | 0.45 | Steep cornea | | + | + | + | | Minimal pigment migration | |
| 75[29] | Mild-Intermediate ZSD | PEX6 p.Q439Gfs3X / p.Q45_G49dup | F | 17 | 0.45 | | | + | | + | | | |
| 76[29] | PEX1 p.G843D/G843D - Mild ZSD | PEX1 p.G843D / G843D | F | 4 | | | | | | | | | |
| 77[30] | Mild-Intermediate ZSD | PEX6 p.L937fs / G437D | F | 4 | 0.2 | | | + | | + | | | |
| 78[30] | Mild-Intermediate ZSD | PEX6 p.L937fs / G437D | M | 6 | 0.25 | | | + | | | | | |
| 79[30] | Mild-Intermediate ZSD | PEX6 p.R142_P143delinsWS / p.R142_P143delinsWS | F | 21 | | | | + | | | | Pre- and retro-equatorial bone spicules, atrophic lesion on the left macula, normal right macula, normal papilla OU | |

Supplementary Table 4.

**Supplementary Table 4 – Continued**

| Reported Patient # | ERG | | VEP | | OCT | | FAF |
|---|---|---|---|---|---|---|---|
| | Scotopic and/or photopic a/b-wave amplitudes | Scotopic and/or photopic a/b-wave latencies | P-100 amplitude (Normal, attenuated (Att.) | Delayed latency of P-100 (Y or N) | Intraretinal cysts (- or +) | Other descriptions | |
| 1[1] | High a-wave amplitude | | Normal | N | | | |
| 2[1] | High a-wave amplitude<br>Small b-wave amplitude | | Normal | N | | | |
| 3[1] | Attenuated a and b wave amplitudes | Delayed a and b waves | Normal | N | | | |
| 4[1] | Attenuated photopic a and b wave amplitudes | | Normal | Y | | | |
| 5[1] | Attenuated photopic and scotopic a and b wave amplitudes | | Normal | Y | | | |
| 6[1] | Attenuated photopic and scotopic a and b wave amplitudes | | Att | N | | | |
| 7[1] | Attenuated a and b wave amplitudes | Delayed a and b waves | Normal | N | | | |
| 8[1] | Attenuated a and b wave amplitudes | Normal a and b wave latencies | Att | Y | | | |
| 9[1] | Attenuated photopic and scotopic responses | Delayed scotopic responses | Normal | N | | | |
| 10[1] | | Normal a and b wave latencies | Normal | N | | | |
| 11[1] | Attenuated a and b wave amplitudes | | Att | N | | | |
| 12[1] | Attenuated a and b wave amplitudes | | Normal | Y | | | |
| 13[1] | Attenuated a and b wave amplitudes | | Normal | N | | | |
| 34[4] | Low amplitude responses (11 µV OD and 9 µV OS) | | Recognizable waveform | Y | | | |
| 37[7] | Absent | | Abnormal | | | | |
| 38[7] | Absent | | Abnormal | | | | |
| 39[7] | Absent | | Abnormal | | | | |
| 40[7] | Absent | | Abnormal | | | | |
| 41[8] | Nonrecordable | | | | | | |

Supplementary Table 4.

| Reported Patient # | ERG | | VEP | | OCT | | FAF |
|---|---|---|---|---|---|---|---|
| | Scotopic and/or photopic a/b-wave amplitudes | Scotopic and/or photopic a/b-wave latencies | P-100 amplitude (Normal, attenuated (Att.) | Delayed latency of P-100 (Y or N) | Intraretinal cysts (- or +) | Other descriptions | |
| 42[8] | Nonrecordable | | | | | | |
| 43[8] | Nonrecordable | | | | | | |
| 44[8] | Attenuated | | | | | | |
| 45[9] | Extinguished | | Att | Y | | | |
| 46[10] | Abolished photopic and 10 µV scotopic responses | | | Y | | | |
| 47[10] | Abolished | | | Y | | | |
| 48[11] | No detectable response | | Abnormal | | | | |
| 49[11] | Abnormal | | Abnormal | | | | |
| 50[11] | Abnormal | | Abnormal | | | | |
| 51[12] | Absent | | Absent | | | | |
| 53[14] | Severe loss of both rod- and cone-driven responses Relatively greater loss of b-wave amplitude at high intensities, leading to decreased b- to a-wave ratio. | | | | - | Outer retinal atrophy with loss of the external limiting membrane and the inner-segment ellipsoid line, hyperreflective opacities suspended in the vitreous, ONL thin, OPL disappeared in places, nerve fiber layer was thickened both nasally and temporally to the fovea OU, severe atrophy of outer retinal structures and pigment epithelium atrophy with nodules of hyper-reflective material on top of the Bruch membrane. | |
| 54[15] | Minimal residual photopic response | Prolonged 30-Hz-flicker times, and scotopic b-waves | | Y | | | |
| 55[15] | Profoundly subnormal scotopic and photopic responses | Prolonged 30-Hz-flicker times | | | | | |
| 56[16] | Absence of photopic and scotopic responses | | | | | | |
| 57[17] | Decreased photopic and scotopic response 95-98% | | | | | | |
| 59[18] | Decreased photopic and scotopic response 95-98% | | | | | | |
| 61[20] | Non-detectable | | | | - | Extreme thinning of fovea with loss of cones in the periphery | Absence of autofluorescence in fovea |

Supplementary Table 4.

| Reported Patient # | ERG | | VEP | | OCT | | FAF |
|---|---|---|---|---|---|---|---|
| | Scotopic and/or photopic a/b-wave amplitudes | Scotopic and/or photopic a/b-wave latencies | P-100 amplitude (Normal, attenuated (Att.) | Delayed latency of P-100 (Y or N) | Intraretinal cysts (- or +) | Other descriptions | |
| 62[21] | Marked subnormal scotopic and minimally subnormal photopic response | | | | + | | Hyperfluorescent disc drusen peripapillary hypofluorescence and hypofluorescent macular mottling |
| 63[22] | Non-detectable scotopic components and no reliable photopic responses | | | | + | Absence of the PR layer in macula OU | |
| 65[24] | Flat response | | | | | | |
| 66[24] | Revealed mild RP | | | | | | |
| 68[25] | Normal | | | | + | Loss of inner/outer segment boundary and RPE thinning | Hyper and hypofluorescent dots corresponding to RPE mottling |
| 69[26] | Rods and cones dysfunction with low voltage traces under white and red flashes stimulation, OD>OS | | Normal | | + | | |
| 70[27] | | | | | - | Depletion of PR in perifoveal area Disruption of the ONL | Hyperfluorescence of perifoveal area |
| 71[27] | | | | | | | Hyperfluorescence of perifoveal area |
| 72[28] | Abnormal photopic and normal scotopic responses | | | | + | | Hyperfluorescence posterior pole sparing fovea |
| 73[28] | Abnormal photopic and normal scotopic responses | | | | - | Depletion of PR and disruption of ONL | Hyperfluorescence paramacular area |
| 74[29] | Decreased amplitude both dark and light adapted response | Delay only in cone flicker | | | + | Macular cystic spaces in 3mm circumference around center of fixation. Retinal thinning. Loss of retinal outer layers OU with foveolar sparing OS. | Hypo-hyperautofluorescence posterior pole and midperiphery. |
| 75[29] | Normal amplitudes photopic and scotopic responses. | Mild delay only in cone flicker | | | + | CMT 927 µm OD, 275 µm OS. Large cystic spaces disrupting macular layers OD, smaller cysts OS | Mostly hyperautofluorescent with a few spots of hypoautofluorescence. |
| 76[29] | | | | | + | CMT 406 µm OD, 466 µm OS | |
| 77[30] | Non-recordable | | Normal | | | Decrease in macular thickness in both eyes, alteration of the photoreceptor layer. | |
| 79[30] | Non-recordable rods and much reduced amplitude for mixed and cones recordings | | | | | | |

Supplementary Table 4.

Supplementary Table 4.

Supplementary Table 4.

29. Varela MD, Jani P, Zein WM, et al. The peroxisomal disorder spectrum and Heimler syndrome: Deep phenotyping and review of the literature. *Am J Med Genet Part C Semin Med Genet*. 2020;184(3):618-630. doi:10.1002/ajmg.c.31823

30. García-García G, Sanchez-Navarro I, Aller E, et al. Exome sequencing identifies PEX6 mutations in three cases diagnosed with Retinitis Pigmentosa and hearing impairment. *Mol Vis*. 2020;26(October 2019):216-225.
