## Supplementary Table 5 for "Peroxisome Biogenesis Disorders in the Zellweger Spectrum: Ophthalmic Findings from a New Natural History Study Cohort and Scoping Literature Review"

**Supplementary Table 5**. **List of Zellweger Spectrum Disorder published case reports and cohort studies and their reported ophthalmic findings**. *The original names of the disorder as stated in the articles were kept. **All visual acuity values shown in this table are LogMAR scores. ±Findings from this article were not included in the Supplementary Table 4 and subsequent data analysis, as we could not assign a disease severity subgroup to the patients because the findings were reported for both severe and mild-intermediate patients together. n = number of patients reported in the article. Abbreviations: ADLs = activities of daily living; CME = cystoid macular edema; DBPD – D-bifunctional protein deficiency; ERG = electroretinogram; ERM = epiretinal membrane; FA = fluorescein angiogram; FAF = fundus autofluorescence; GCL = ganglion cell layer; ILM = internal limiting membrane; INL = inner nuclear layer; IOP = intraocular pressure; NGS = next-generation sequencing; OCT = optical coherence tomography; OD = right eye; ON = optic nerve; ONL = outer nuclear layer; OPL = outer plexiform layer; OS: left eye; OU = both eyes; PBD = Peroxisome biogenesis disorder; PR = photoreceptor; RNFL = retinal nerve fiber layer; RP = retinitis pigmentosa; RPE = retinal pigment epithelium; VA = visual acuity; VEP = visual evoked potential; VF = Visual field; ZSD = Zellweger Spectrum Disorder.

| **Author (Year)** | **Type of Study** | **n** | **Disease severity subgroup** | **Description*** | **Endpoint measures** | | | | **Clinical and pathological findings** |
|---|---|---|---|---|---|---|---|---|---|
| | | | | | **ERG** | **VEP** | **FAF** | **OCT** | |
| Noguer et al. (2010)[1] | Case series | 23 | ZSD (all spectrum): 20 Mild-Intermediate, 2 Severe | Visual data obtained in patients under DHA-EE supplementation | X | X | | | - VA: stable in all patients<br>- Fundus: stable retinal exam in 22/23 patients<br>- Nystagmus: disappeared in all patients<br>- ERG and VEP: general improvement in all patients |
| Berendse et al. (2016)[2] | Retrospective cohort | 19 | ZSD (all spectrum) | Clinical manifestations and some ocular findings | | | | | - 78% of patients had VA of 0.7 or worse**<br>- Mean VA was 0.85 (ranging from 1.3 to 0.5)<br>- 100% had nyctalopia and retinopathy |
| Haddad et al. (1976)[3] | Case report | 1 | Severe ZSD | Ocular pathologic findings in an autopsy of a neonate with "Cerebro-Hepato-Renal Syndrome of Zellweger" | | | | | Both eyes:<br><br>- Cornea: thickened and showed paracentral iridocorneal adhesions, corresponding to stromal edema with thin and disorganized lamellae; Descemet membrane was thin<br>- Anterior segment: shallow with developed Schlemm canals but focal ciliary body necrosis<br>- Lens: posterior subcapsular cataracts<br>- Optic disc: hypercellular with loss of nerve fibers<br>- Fundus: thinning and PR loss in the posterior pole and perimacular area |

Supplementary Table 5.

| Author (Year) | Type of Study | n | Disease severity subgroup | Description* | Endpoint measures ERG | VEP | FAF | OCT | Clinical and pathological findings |
|---|---|---|---|---|---|---|---|---|---|
| Garner et al. (1982)[4] | Case report | 1 | Severe ZSD | Ocular findings and ocular pathological findings in a patient with "Cerebro-hepato-renal (Zellweger's) syndrome" | X | X | | | - VA: LP<br>- Jerk nystagmus on lateral gaze<br>- Normal anterior segments<br>- Optic disc: normal<br>- Fundus: Retinal arteries attenuated. Red lesion 1/3 of disc diameter in macula OD. Deep pigment clumping surrounded by rim of depigmentation in macula OS.<br>- ERG: Low amplitude response obtained (11 µV OD and 9 µV OS)<br>- VEP: Recognizable waveform, delayed latency<br>Pathological findings:<br>- Atrophic flattening of the RPE was widespread, with focal deficiencies and migration of occasional cells away from their attachment to Bruch's membrane<br>- Electron microscopy: Almost total loss of the PR outer segments |
| Glasgow et al. (1987)[5] | Case report | 1 | Severe ZSD | Pathologic findings of two eyes at autopsy of a 15-month old with "NALD" | | | | | - Pupils at birth: reactive to light<br>- Vitreous: numerous white vitreous opacities corresponding to PAS-positive macrophages<br>- Optic disc: diffuse atrophy of the GCL and RNFL<br>- Fundus: focal retinal pigmentation in the temporal periphery corresponding to intracellular and extracellular multi-layered pigment; marked PR cell degeneration in the periphery and within the macula |
| Colburn et al. (2009)[6] | Case report | 1 | Severe ZSD | Description of a vitamin A deficiency-associated corneal ulcer in a patient with "Zellweger syndrome" | | | | | - Age-appropriate fix-and-follow behavior but severe photophobia<br>- IOP: 20 mm Hg OD, 35 mm Hg OS<br>- Cornea OD: dry, granular epithelium and a diffusely cloudy appearance<br>- Cornea OS: 4-5 mm central corneal ulcer with raised and infiltrated edges, secondary to vitamin A deficiency<br>- Anterior chamber OS: 2-3 mm hypopyon<br>- Fundus: normal OU<br>- Cultures grew *Streptococcus constellatus*<br>- Serum retinol concentration 0.05mg/L (normal 0.20 – 0.50)<br>Treatment and evolution<br>- Fortified vancomycin and cefazolin every hour OS with no improvement<br>- Vitamin A oral supplementation (17,200 units daily x 7 days) with resolution of the ulcer OS and cloudy cornea OD<br>- The corneal ulcer improved with vitamin A supplementation |

Supplementary Table 5.

| Author (Year) | Type of Study | n | Disease severity subgroup | Description* | Endpoint measures | | | | Clinical and pathological findings |
|---|---|---|---|---|---|---|---|---|---|
| | | | | | ERG | VEP | FAF | OCT | |
| Hittner et al. (1981)[7] | Case series | 4 | Severe ZSD | Ocular findings in patients with "Zellweger syndrome" | X | X | | | - Zonular cataracts in all patients<br>- Macular degeneration: patients 3 and 4<br>- Retinal deposits: moderate (patient 3) and severe (patient 4)<br>- Optic disc: atrophied in patients 3 and 4<br>- ERG: absent in all patients, VEP: abnormal in all patients |
| Govaerts et al. (1982)[8] ± | Case series | 16 | Severe and Mild-Intermediate ZSD | Ocular signs in patients with "Cerebro-Hepato-Renal Syndrome of Zellweger" | | | | | - Cataracts: 2/6 patients (33%)<br>- Peripheral retinal pigment clumping: 4/11 (36%)<br>- Optic disc pallor: 8/11 patients (73%)<br>- Nystagmus: 10/11 patients (91%) |
| Lambert et al. (1989)[9] | Case report | 4 | Severe and Mild-Intermediate ZSD | Ocular examinations in 2 patients with "Zellweger syndrome" and "Infantile Refsum's disease" previously diagnosed as Leber congenital amaurosis (LCA) | X | | | | Patients 1 and 2 (Severe ZSD):<br>- Nystagmus<br>- Fundus: Diffuse pigmentary changes<br>- ERG: Nonrecordable<br>Patient 3 (Mild ZSD):<br>- Nystagmus<br>- Fundus: Diffuse pigmentary changes<br>- ERG: Nonrecordable<br>Patient 4 (Mild ZSD):<br>- Nystagmus<br>- Cataracts<br>- ERG: Attenuated |
| Stanescu-Segall et al. (1989)[10] | Case report | 1 | Mild-Intermediate ZSD | Retinopathy in a patient with "cerebro-hepatorenal syndrome of Zellweger" | X | X | | | - Visually inattentive<br>- Pupils: mildly dilated, slow light reflex<br>- Anterior segment: normal<br>- Optic disc: pale with exuberant glial tissue<br>- Fundus: attenuation of retinal vessels and pigmentary clumping<br>- ERG: extinguished, VEP: delayed latency and low amplitude |
| Stanescu-Segall (1996)[11] | Case report | 3 | Severe ZSD | Retinopathy in patients with "cerebro-hepatorenal syndrome of Zellweger" | X | X | | | - Optic disc: pallor in patients 1 and 2<br>- Fundus: attenuation of retinal vessels (patient 1), retinal pigmentary mottling in all patients<br>- ERG: abolished photopic and 10 microvolt scotopic in patient 1, abolished in patients 2 and 3. VEP: prolonged latency in all patients |

Supplementary Table 5.

| Author (Year) | Type of Study | n | Disease severity subgroup | Description* | Endpoint measures | | | | Clinical and pathological findings |
|---|---|---|---|---|---|---|---|---|---|
| | | | | | ERG | VEP | FAF | OCT | |
| Lyons et al. (2004)[12] | Case report | 3 | Mild-Intermediate ZSD | Leopard spot retinal pigmentation in patients with “NALD”. | X | X | | | - Visual inattention in all patients<br>- Pupils: sluggish reflexes in patient 2<br>- Cornea: subtly clouding in patient 2<br>- Fundus: peripheral leopard spot pigmentary retinopathy in all patients and later, retinal blood vessel attenuation in patient 2<br>- Nystagmus: horizontal in patients 2 and 3<br>- ERG and VEP: abnormal in all patients |
| Michelakakis et al. (2004)[13] | Case report | 1 | Mild-Intermediate ZSD | Case of PEX1 deficiency diagnosed as LCA | X÷ | X÷ | | | - Optic disc: papilledema OU<br>- Fundus: tapetoretinal degeneration (salt and pepper)<br>- ERG and VEP: absence of any response. |
| O' Bryhim et al. (2018)[14] | Case report | 1 | Intermediate ZSD | Novel retinal findings in a patient with PBD | | | | | - Ability to fixate, follow and maintain fixation OU<br>- Fundus: large chorioretinal folds with fibrosis OD and ERM with peripheral pigmentary changes OS<br>- Optic disc: normal OU<br>- Strabismus: large angle intermittent exotropia<br>- Latent nystagmus |
| Courtney et al. (2013)[15] | Case report | 1 | Mild-Intermediate ZSD | OCT and ERG findings in a patient with Neonatal Adrenoleukodystrophy | X | | | X | - VA F+F OD and OS<br>- IOP normal<br>- Refraction: -3.50 +0.50 x 90 OU<br>- Clear corneas, lamellar/sutural cataracts OU<br>- Fundus: Normal ON and vessels, macula dull without foveal light reflex, chorioretinal atrophy with pigmentary changes in leopard-spot pattern through midperiphery.<br>- ERG: Severe loss of both rod- and cone-driven responses. Relatively greater loss of b-wave amplitude at high intensities, leading to decreased b- to a-wave ratio.<br>- OCT: Outer retinal atrophy with loss of the external limiting membrane and the inner-segment ellipsoid line, hyperreflective opacities suspended in the vitreous, ONL thin, OPL disappeared in places, nerve fiber layer was thickened both nasally and temporally to the fovea OU, severe atrophy of outer retinal structures and pigment epithelium atrophy with nodules of hyper-reflectivematerial on top of the Bruch membrane.<br>- |

Supplementary Table 5.

| Author (Year) | Type of Study | n | Disease severity subgroup | Description* | Endpoint measures ERG | VEP | FAF | OCT | Clinical and pathological findings |
|---|---|---|---|---|---|---|---|---|---|
| Weleber et al. (1984)[16] | Case report | 2 | Mild-Intermediate ZSD | Ocular findings in patients with "Infantile Phytanic acid storage disease" | X÷ | X | | | Patient 1:<br>- Refraction: +2.00 +1.00 x 90 OU<br>- Fundus: pigmentary epithelium mottling OU with possibility of tapetoretinal degeneration<br>- Optic disc: pallor OU<br>- Nystagmus<br>- ERG: minimal residual photopic cone response, prolonged 30-Hz-flicker times, prolonged scotopic b-waves, VEP: prolonged latency OD<br>- FA: marked loss of RPE and choriocapillaris with attenuation of retinal vessels<br>Patient 2:<br>- Age-appropriate visual fixation<br>- Refraction: +1.25 sphere OU<br>- Anterior segment: normal<br>- Fundus: diffusely hypopigmented macula with mottling of the RPE and wrinkling of the ILM<br>- Optic disc: normal<br>- Nystagmus: absent<br>- Strabismus: absent<br>- ERG: profoundly subnormal cone and rod responses and prolonged 30-Hz-flicker times |
| Ek et al. (1986)[17] | Case report | 1 | Mild-Intermediate ZSD | Visual manifestations in a patient with LCA | X | | | | - Absence of visual contact<br>- Pupils: dilated, no response to light<br>- IOP: normal<br>- Anterior segment: normal<br>- Fundus: no foveal reflex, narrow retinal vessels, hyperpigmentation of the maculae OU with more abundant pigmentation in the periphery<br>- Optic disc: grey and edematous papillae<br>- ERG: absence of photopic and scotopic responses |
| Torvik et al. (1988)[18] | Case report | 1 | Mild-Intermediate ZSD | Pathologic findings in an autopsy of a patient with "Infantile Refsum disease" | | | | | - Fundus: marked atrophy of all retinal layers throughout the retinal circumference<br>- Loss of rods and cones without near complete disappearance of the ONL and INL<br>- Scattered pigmentary retinal clumps |

Supplementary Table 5.

| Author (Year) | Type of Study | n | Disease severity subgroup | Description* | Endpoint measures | | | | Clinical and pathological findings |
|---|---|---|---|---|---|---|---|---|---|
| | | | | | ERG | VEP | FAF | OCT | |
| Folz et al. (1991)[19] | Review and case reports | 2 | Mild-Intermediate ZSD | Ocular descriptions in patients with "NALD". | X | | | | - VA: 1.00 OU in both patients<br>- Pupils: moderately reactive but slugging to light in both patients<br>- Fundus: clumped retinal pigment in the mid-periphery with perifoveal depigmentation in both patients<br>- Optic disc: waxy pallor with marked attenuation of the retinal vessels in both patients<br>- Strabismus : concomitant 30 prism diopter exotropia OD (patient 1), no deviation (patient 2)<br>- Nystagmus: absent in both patients<br>- ERG: decreased cone and rod response 95 to 98% in both patients |
| Berman (2013)[20] | Case report | 1 | Mild-Intermediate ZSD | Ophthalmological exam in a *PEX1* patient | | | | | - Visually inattentive<br>- Iris: unpigmented but dark in the periphery<br>- Pupils: poor constriction<br>- IOP: 20 mmHg OU<br>- Fundus: hypoplastic choroid with moderate vascularization and multiple small dark pigments present at the equatorial regions<br>- Optic disc: poor vascularization<br>- Strabismus: intermittent esotropia<br>- Nystagmus: jerk up-nystagmus |
| Majewski et al. (2011)[21] | Case report | 1 | Mild ZSD | Novel ocular findings in a patient with PBD | X | | X | X | - VA: 1.30 OU<br>- VF: central scotomas<br>- Pupils: amaurotic pupils<br>- Fundus: marked pigmentary maculopathy with a relatively intact perifoveal retina and RPE atrophy<br>- Optic disc OS: optic disc pallor with drusen and narrow vessels<br>- Nystagmus: present<br>- ERGs: non-detectable<br>- FAF: absence of autofluorescence in the fovea<br>- OCT: extreme thinning of the fovea with loss of cones in the periphery |

Supplementary Table 5.

| Author (Year) | Type of Study | n | Disease severity subgroup | Description* | Endpoint measures | | | | Clinical and pathological findings |
|---|---|---|---|---|---|---|---|---|---|
| | | | | | ERG | VEP | FAF | OCT | |
| Pakzad-Vaezi et al. (2014)[22] | Case report | 1 | Mild ZSD | Ocular findings in a patient with “Infantile Refsum disease” | X | | X | X | - VA: 0.70 OD, 0.60 OS<br>- Goldmann VF: generalized constriction with a few mid-peripheral islands of preserved vision<br>- Refraction: low myopia OS<br>- Lens: early subcapsular cataract OS<br>- Fundus: non-corpuscular pigmentary degeneration and attenuated retinal vasculature<br>- Optic disc: prominent optic nerve head drusen<br>- Strabismus: 35-diopter esotropia OD<br>- ERG: marked subnormal scotopic response and minimally subnormal photopic response<br>- FAF: hyperfluorescent disc drusen, peripapillary hypofluorescence and hypoluorescent macular mottling<br>- OCT: cystoid macular edema (CME) |
| Ventura et al. (2016)[23] | Case report | 1 | Mild ZSD | Ocular findings in a PBD patient diagnosed as “Usher syndrome” | X | | | X | - VA: 0.65 OD, 0.72 OS<br>- Fundus: pigmentary retinopathy<br>- Optic disc: optic nerve hypoplasia<br>- ERG: non-detectable rod components and no reliable cone responses<br>- OCT: CME and absence of the PR layer in the macula OU |
| Neuhaus et al. (2017)[24] | Case report | 1 | Mild-Intermediate ZSD | Ocular findings in a patient diagnosed with “Heimler Syndrome” through NGS | | | | | - Significant visual loss<br>- Retinitis punctata albescens with macular involvement |
| Raas-Rothschild et al (2002)[25] | Case report | 3 | Severe ZSD and Mild-Intermediate ZSD | Ocular findings in one severe PBD infant and in mild affected parents resembling “Usher Syndrome” | X | X | | | Patient 1 (Severe ZSD infant):<br>- Peripheral pigmentary changes consistent with RP<br>- Small optic discs<br>- ERG: Flat response<br>- VEP: Results compatible with RP<br>Patient 2 (Mild ZSD parent):<br>- Night blindness<br>- VF: Restriction of peripheral VF to 100º<br>Patient 3 (Mild ZSD parent):<br>- Ophthalmologic evaluation and ERG revealed RP |

Supplementary Table 5.

| Author (Year) | Type of Study | n | Disease severity subgroup | Description* | Endpoint measures | | | | Clinical and pathological findings |
|---|---|---|---|---|---|---|---|---|---|
| | | | | | ERG | VEP | FAF | OCT | |
| Lima et al. (2011)[26] | Case report | 1 | Mild-Intermediate ZSD | Ocular findings in a patient with "Heimler Syndrome"÷ | X | | X | X | - VA: 1.00 OD, 0.30 OS<br>- FA: mottling of the RPE in the posterior pole and periphery<br>- FAF: hyper and hypoautofluorescent dots corresponding to RPE mottling<br>- OCT: loss of the inner/outer segments boundary and RPE thinning<br>- ERG: no generalized rod-cone dysfunction |
| Ratbi et al. (2016)[27] | Case report | 1 | Mild-Intermediate ZSD÷ | Ocular findings in a patient with "Heimler Syndrome"÷ | X | X | | X | - VA: Rapid and important decrease<br>- Nyctalopia<br>- Anterior segment: normal<br>- IOP: normal<br>- Nystagmus: Rotatory<br>- Fundus: decreased macular reflection with edema, pigmented deposits, slender and rigid retinal arteries and a pale yellow waxy papilla<br>- ERG: Rods and cones dysfunction with low voltage traces under white and red flashes stimulation, OD>OS, VEP: Normal<br>- OCT: CME OS.<br>- Treatment with acetazolamide, intravitreal triamcinolone and intravitreal anti-VEGF without any amelioration of the macular edema. |
| Smith et al. (2016)[27] | Case report | 2 | Mild-Intermediate ZSD | Ocular findings in patients with "Heimler Syndrome" | | | X | X | - Retinal dystrophy (both patients)<br>- Fundus: pigmentary maculopathy and mild retinal vascular attenuation (OD, patient 1)<br>- OCT: depletion of PR in the perifoveal area and disruption of the ONL (OD, patient 1)<br>- FAF: hyperfluorescence of the perifoveal area (both patients) |

Supplementary Table 5.

| Author (Year) | Type of Study | n | Disease severity subgroup | Description* | Endpoint measures ERG | VEP | FAF | OCT | Clinical and pathological findings |
|---|---|---|---|---|---|---|---|---|---|
| Wangtiraumnuay et al. (2018)[28] | Case report | 2 | Mild-Intermediate ZSD÷ | Ocular findings in patients with “Heimler Syndrome” | X÷ | | X | X | Patient 1 :<br>- VA: 0.8 OD, 2.3 OS<br>- Refraction +1.50 OD, +2.00 OS<br>- Goldman VF: Normal<br>- Cornea and iris: normal<br>- Lens: Cortical flecks OU<br>- Fundus: Peripheral hyperpigmentation and scattered round hyperpigmented lesions<br>- Optic disc and retinal vessels: normal<br>- ERG: Abnormal cones and normal rods<br>- FAF: hyperfluorescent deposits throughout posterior pole sparing fovea<br>- OCT: Macular intraretinal cystoid spaces<br>Patient 2 :<br>- VA 0.00 OU<br>- Refraction +1.00 OD, +0.50 OS<br>- Goldman VF: Normal<br>- Iris: Single small focus of transillumination<br>- Cornea and lens: normal<br>- Lens: Cortical flecks OU<br>- Fundus: pigmentary maculopathy and mid periphery mottling<br>- Optic disc and retinal vessels: normal<br>- ERG: Abnormal cones and normal rods<br>- FAF: hyperfluorescent deposits throughout paramacular area<br>- OCT: depletion of PR and disruption of ONL |
| Varela et al. (2020)[29] | Review and case reports | 3 | Mild ZSD and Mild-Intermediate ZSD | Ocular, audiological and dental findings in two patients with “Heimler Syndrome” and one patient with “Mild-intermediate Peroxisomal Disorder Spectrum” and literature review of mild peroxisomal disorders | X | | X | X | Patient 1:<br>- VA: 0.8 OD, 0.1 OS<br>- Goldman VF: Preserved peripheral vision (V4e isopter), paracentral temporal scotoma (I4e isopter), functional central scotoma (I1e isopter).<br>- Steep cornea (K1 48.98, K2 47.8)<br>- IOP: Normal<br>- Axial length: Shorter than average (20.3 OD, 20.34 OS)<br>- Deficiency in color discrimination OD<br>- Fundus: Retinal vascular attenuation, mottled RPE, minimal pigment migration<br>- Optic disc pallor |

Supplementary Table 5.

| Author (Year) | Type of Study | n | Disease severity subgroup | Description* | Endpoint measures ERG | VEP | FAF | OCT | Clinical and pathological findings |
|---|---|---|---|---|---|---|---|---|---|
| | | | | | | | | | - ERG: Decreased amplitude both dark and light adapted response, delay only in cone flicker.<br>- FAF: Hypo-hyperautofluorescence posterior pole and midperiphery.<br>- OCT: Macular cystic spaces in 3mm circumference around center of fixation. Retinal thinning (CMT 123 µm OD, 198 µm OS). Loss of retinal outer layers OU with foveolar sparing OS.<br>Patient 2:<br>- VA: 0.9 OD, 0.0 OS<br>- Goldman VF: Functional central scotoma OU, preserved periphery<br>- IOP: Normal<br>- Deficiency in color discrimination OD<br>- Fundus: Mottled posterior pole and mid periphery, mild vascular attenuation.<br>- ERG: Normal amplitudes photopic and scotopic responses. Mild delay only in cone flicker.<br>- FAF: Mostly hyperautofluorescent with a few spots of hypoautofluorescence.<br>- OCT: CMT 927 µm OD, 275 µm OS. Large cystic spaces disrupting macular layers OD, smaller cysts OS.<br>Patient 3 (Diagnosed with ZSD and Marfan syndrome)<br>- Axial length and IOP: Normal<br>- Ishihara color test 14/16 plates read<br>- OCT: CMT 406 µm OD, 466 µm OS. Cystic spaces OU. |
| García-García et al. (2020)[30] | Case report | 3 | Mild-Intermediate ZSD | Audiological and ocular findings in patients with *PEX6* mutations found by whole exome sequencing (WES) | X | X | | X | Patient 1:<br>- VA: 0.3 OD, 0.1 OS<br>- Fundus: Abnormal retinal pigmentation with peripheral pigment accumulation, Arterial narrowing.<br>- ERG: Non-recordable<br>- VEP: Normal<br>- OCT: Decrease in macular thickness in both eyes, alteration of the photoreceptor layer.<br>Patient 2 (Patient 1's sibling):<br>- VA: 0.4 OD, 0.1 OS<br>- Fundus: Peripheral temporal pigment deposits suggesting RP.<br>Patient 3: |

Supplementary Table 5.

| Author (Year) | Type of Study | n | Disease severity subgroup | Description* | Endpoint measures | | | | Clinical and pathological findings |
|---|---|---|---|---|---|---|---|---|---|
| | | | | | ERG | VEP | FAF | OCT | |
| | | | | | | | | | - Night blindness<br>- VF constriction<br>- Fundus: Pre- and retro-equatorial bone spicules, atrophic lesion on the left macula, normal right macula, normal papilla OU.<br>- ERG: Non-recordable rods and much reduced amplitude for mixed and cones (flash and flicker ERG recordings). |

Supplementary Table 5.

Supplementary Table 5.

Supplementary Table 5.


29. Varela MD, Jani P, Zein WM, et al. The peroxisomal disorder spectrum and Heimler syndrome: Deep phenotyping and review of the literature. *Am J Med Genet Part C Semin Med Genet*. 2020;184(3):618-630. doi:10.1002/ajmg.c.31823

30. García-García G, Sanchez-Navarro I, Aller E, et al. Exome sequencing identifies PEX6 mutations in three cases diagnosed with Retinitis Pigmentosa and hearing impairment. *Mol Vis*. 2020;26(October 2019):216-225.
